# Heparin for Moderately Ill Patients with Covid-19

**DOI:** 10.1101/2021.07.08.21259351

**Authors:** Michelle Sholzberg, Grace H. Tang, Hassan Rahhal, Musaad AlHamzah, Lisa Baumann Kreuziger, Fionnuala Ní Áinle, Faris Alomran, Khalid Alayed, Mohammed Alsheef, Fahad AlSumait, Carlos Eduardo Pompilio, Catherine Sperlich, Sabrena Tangri, Terence Tang, Peter Jaksa, Deepa Suryanarayan, Mozah Almarshoodi, Lana Castellucci, Paula D. James, David Lillicrap, Marc Carrier, Andrew Beckett, Christos Colovos, Jai Jayakar, Marie-Pier Arsenault, Cynthia Wu, Karine Doyon, E. Roseann Andreou, Vera Dounaevskaia, Eric K. Tseng, Gloria Lim, Michael Fralick, Saskia Middeldorp, Agnes Y.Y. Lee, Fei Zuo, Bruno R. da Costa, Kevin E. Thorpe, Elnara Márcia Negri, Mary Cushman, Peter Jüni, the RAPID Trial investigators

## Abstract

**Background:** Heparin, in addition to its anticoagulant properties, has anti-inflammatory and potential anti-viral effects, and may improve endothelial function in patients with Covid-19. Early initiation of therapeutic heparin could decrease the thrombo-inflammatory process, and reduce the risk of critical illness or death.

**Methods:** We randomly assigned moderately ill hospitalized ward patients admitted for Covid-19 with elevated D-dimer level to therapeutic or prophylactic heparin. The primary outcome was a composite of death, invasive mechanical ventilation, non-invasive mechanical ventilation or ICU admission. Safety outcomes included major bleeding. Analysis was by intention-to-treat.

**Results:** At 28 days, the primary composite outcome occurred in 37 of 228 patients (16.2%) assigned to therapeutic heparin, and 52 of 237 patients (21.9%) assigned to prophylactic heparin (odds ratio, 0.69; 95% confidence interval [CI], 0.43 to 1.10; p=0.12). Four patients (1.8%) assigned to therapeutic heparin died compared with 18 patients (7.6%) assigned to prophylactic heparin (odds ratio, 0.22; 95%-CI, 0.07 to 0.65). The composite of all-cause mortality or any mechanical ventilation occurred in 23 (10.1%) in the therapeutic heparin group and 38 (16.0%) in the prophylactic heparin group (odds ratio, 0.59; 95%-CI, 0.34 to 1.02). Major bleeding occurred in 2 patients (0.9%) with therapeutic heparin and 4 patients (1.7%) with prophylactic heparin (odds ratio, 0.52; 95%-CI, 0.09 to 2.85).

**Conclusions:** In moderately ill ward patients with Covid-19 and elevated D-dimer level, therapeutic heparin did not significantly reduce the primary outcome but decreased the odds of death at 28 days.

Trial registration numbers: NCT04362085; NCT04444700

## INTRODUCTION

The most common cause of clinical deterioration of patients hospitalized for Covid-19 is hypoxic respiratory failure.^1^ Pulmonary vascular dysfunction and microvascular thrombosis likely contribute to respiratory compromise,^2–6^ which in turn may be caused by a thrombo-inflammatory state also referred to as Covid-19 coagulopathy.^2,3,7–9^ Elevated D-dimer levels, as marker of coagulopathy, and hypoxia are associated with poor prognosis.^2,3,7–10^

Heparin, in addition to its anticoagulant properties, has anti-inflammatory and potential anti-viral effects, and may improve endothelial function.^2,11–15^ Early initiation of therapeutic heparin could therefore decrease the thrombo-inflammatory process, and reduce the risk of critical illness or death.^2,4,16–18^ Randomized trials indicated that therapeutic heparin anticoagulation therefore may be beneficial in moderately,^19^ but not critically ill^20^ patients with Covid-19, suggesting that the time of initiation of therapeutic heparin is indeed important. The Therapeutic Anticoagulation versus Standard Care as a Rapid Response to the COVID-19 Pandemic (RAPID) trial was designed to determine if therapeutic heparin is superior to prophylactic heparin in moderately ill ward patients with Covid-19 and elevated D-dimer level in decreasing the composite of intensive care unit (ICU) admission, mechanical ventilation, or death.

## METHODS

### Trial Design and Oversight

The RAPID trial was an investigator-initiated, parallel, pragmatic, adaptive multi-center, open-label randomized controlled trial conducted at 28 sites in 6 countries. For administrative reasons, the protocol in Brazil was registered separately, but prospectively harmonized (see Supplementary Appendix). The trial was designed by the co-principal investigators and supported by St. Michael’s Hospital Foundation and the Canadian Institute for Military and Veteran Health Research and multiple participating hospitals (see Supplementary Appendix).^21^ The funders had no role in trial design, conduct, collection, management, analysis or interpretation of data, or in preparation or review of the manuscript or the approval of the manuscript for submission. An independent data and safety monitoring board oversaw the trial (see Supplementary Appendix). Authorized research ethics committees approved the trial at all participating sites. The authors vouch for completeness and accuracy of the data and the fidelity of the trial to the protocol.^21^

### Participants

Patients were eligible if they were admitted to hospital wards for Covid-19 with laboratory confirmed SARS-CoV-2 infection and elevated D-dimer levels within the first 5 days of admission. D-dimer levels were required to be above the upper limit of normal (ULN) of the local hospital in the presence of an oxygen saturation ≤93% on room air, or ≥2 times the ULN irrespective of oxygen saturation. Participants were excluded if they had substantial bleeding risks, an absolute indication for or any contraindication against heparin anticoagulation based on care team judgment, were pregnant or if they had already met, or would imminently meet any component of the primary outcome. Full eligibility criteria are provided in the Supplementary Appendix. Written informed consent was obtained from all participants or their legal representatives.

### Randomization and Interventions

We used web-based central randomization with a computer-generated random sequence with variable block sizes stratified by site and age (≤65 versus >65 years) to assign patients in a 1:1 ratio to therapeutic or prophylactic heparin. Patients allocated to therapeutic heparin received therapeutic doses of low molecular weight heparin (LMWH) or unfractionated heparin (UFH), patients allocated to prophylactic heparin received dose-capped prophylactic subcutaneous heparin (LMWH or UFH) adjusted for body mass index and creatinine clearance (see Supplementary Appendix for dosing).

Study treatment was started within 24 hours after randomization, and continued until the first of hospital discharge, day 28, study withdrawal or death (Fig. S1). If a participant was admitted to ICU, continuation of the allocated treatment was recommended.

### Outcomes and Follow-up

The primary outcome was a composite of ICU admission, non-invasive (bilevel or continuous positive airway pressure) or invasive mechanical ventilation, or death up to 28 days. Secondary outcomes included all-cause death; the composite of any mechanical ventilation or all-cause death; ICU admission or all-cause death; ventilator-free days alive; organ support-free days alive; ICU-free days alive; hospital-free days alive (see Supplementary Appendix); renal replacement therapy; venous thromboembolism; arterial thromboembolism; and D-dimer level at 2 days ± 24 hours post-randomization (see Supplementary Appendix for detailed definitions). The following components of the primary composite were not included in the protocol, but pre-specified as secondary outcomes in the statistical analysis plan: invasive mechanical ventilation; composite of invasive or non-invasive mechanical ventilation; ICU admission. Pre-specified safety outcomes included: major bleeding as defined by the International Society on Thrombosis and Haemostasis; ^22^ red blood cell transfusion (≥1 unit); transfusion of hemostatic blood components or products; and heparin-induced thrombocytopenia. An independent event-adjudication committee, which consisted of clinicians who were unaware of the treatment assignments, adjudicated the components of the primary outcome, bleeding and thrombotic events.

### Statistical Analysis

We estimated that 231 patients per group would provide 90% power to detect a 15% absolute risk difference, from 50% in the control group to 35% in the experimental group, at a two-sided alpha level of 0.048 accounting for two planned interim analyses at 25% and 75% of the originally planned sample size.^2,16,21^ At 75%, we performed a conditional power analysis. The protocol pre-specified that the sample size would be increased if the conditional power was between 60 and 80%.^21^ As the conditional power was below 60%, the data safety and monitoring board recommended not to increase the sample size and complete recruitment as originally planned.

The primary analysis of the primary composite outcome was based on the intention-to-treat (ITT) principle using logistic regression; a Chi-squared test of independence was conducted to obtain the two-sided p-value.^23^ Participants who did not have a 28-day assessment, but discharged from hospital alive prior to day 28, were assumed to be event-free up to day 28. Pre-specified subgroup analyses of the primary outcome accompanied by tests of interaction were done for age, sex, body mass index, time from Covid-19 symptom onset, diabetes mellitus, coronary artery disease, hypertension, and race/ethnicity, D-dimer level, use of systemic corticosteroid and geographical region. Pre-specified sensitivity analyses of the primary outcome excluded participants who did not have a 28-day assessment, and included only participants from the per-protocol cohort. The per-protocol cohort was all eligible participants who received their intervention as randomly allocated during the first 48 hours after randomization, and excluded participants who did not satisfy all eligibility criteria, did not receive their allocated treatment or did not have follow-up until day 28, death, or other primary composite outcome component. Since randomization was stratified by age an additional logistic regression model was fit to analyse the primary outcome controlling for age.

Binary secondary outcomes were analyzed using logistic regression. Ventilator-free days, organ support-free days, ICU-free days and hospital-free days alive were analysed using ordinal logistic regression; death up to 28 days was assigned the worst outcome (a value of −1) in these analyses.^24^ The post-treatment D-dimer was compared using linear regression adjusted for baseline. Since D-dimer assays differed across sites, D-dimer levels were analyzed as the logarithm of D-dimer x ULN by taking the natural logarithm of the ratio of the actual d-dimer value divided by the ULN for the assay used. Analyses of secondary outcomes were considered exploratory so were not adjusted for multiple comparisons; the widths of 95% confidence intervals for secondary outcomes should not be used for inferences about treatment effects. An extended description of the statistical methods is provided in the Supplementary Appendix.

Mantel-Haenszel fixed-effect meta-analyses of odds ratios were done for outcomes reported in available trials of therapeutic heparin compared with usual care using lower doses of heparin in hospitalized patients with Covid-19. Organ support-free days in the RAPID trial were recalculated for an observation time of 21 days, in accordance with the primary outcome definition used in the other trials.^19,20^ Analyses were done separately for moderately ill ward patients and severely ill ICU patients, using a chi-squared test to estimate p-values for interaction between treatment and severity of illness. The variance attributed to pooled results reflects only sampling error due to the play of chance at randomization. Homogeneity of odds ratios is not required for fixed-effect pooled odds ratios to be informative.^25^

## RESULTS

From May 29, 2020 through April 12, 2021, a total of 3975 patients were screened, 465 were randomized: 228 were assigned to therapeutic heparin group and 237 to prophylactic heparin, and 222 (97.4%) and 232 (97.9%) received treatment as allocated during the first 48 hours after randomization (Fig. 1).

**Figure 1.**
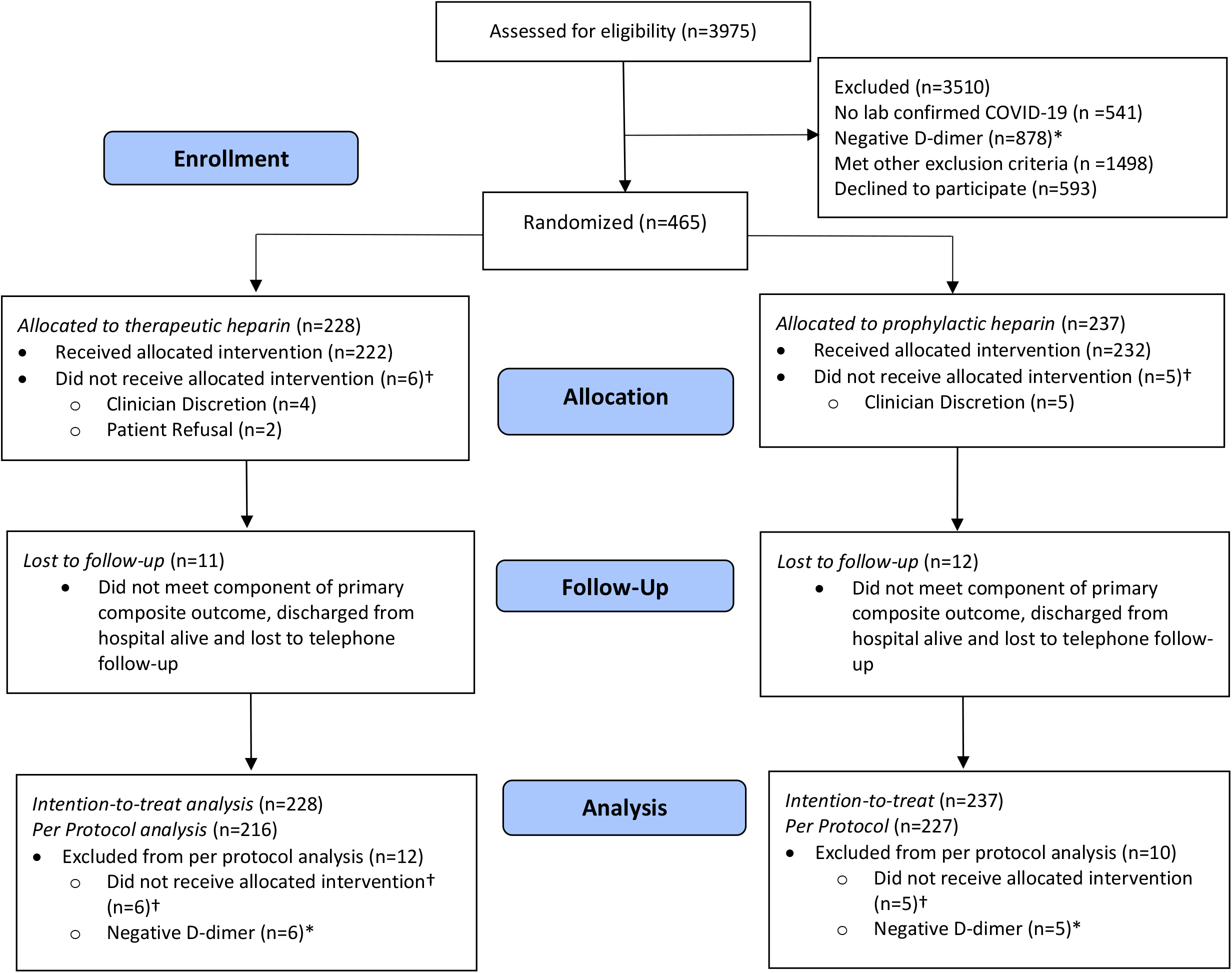
Screening, Enrollment, Randomization, and Inclusion in Analysis. *Negative D-dimer is either D-dimer <2 times upper limit of normal without hypoxia, or D-dimer < upper limit of normal with hypoxia; 6 patients in the therapeutic heparin group and 5 prophylactic heparin group did not meet eligibility criteria pertaining to D-dimer at the time of randomization due to a delay in protocol harmonization with Brazil. †Did not receive allocated intervention within the first 48 hours post-randomization without clear clinical indication.

The mean age was 60 years; 264 participants (56.8%) were men and the mean body mass index was 30.3. Baseline D-dimer was 2.3-fold above the ULN and mean creatinine was 85.2 µmol/L. Six patients in the therapeutic and five in the prophylactic heparin group had D-dimer levels below the ULN at baseline. Baseline demographic and clinical characteristics were similar between treatment arms (Table 1). The mean duration from symptom onset to hospitalization was 7.1 days, and the mean duration from hospitalization to randomization was 1.4 days. The mean treatment duration was 6.5 and 6.3 days in therapeutic and prophylactic heparin groups, respectively (Table S1). LMWH was prescribed in 224 (98.2%) of patients assigned to therapeutic and 222 (93.7%) patients assigned to prophylactic heparin. Concomitant treatments (systemic corticosteroids, remdesivir, and tocilizumab) were balanced between treatment arms for pre- and post-randomization periods combined (Table S2).

**Table 1.** Characteristics of the Patients at Baseline, According to Treatment Assignment*.

| **Characteristic** | **Therapeutic**<br>**Heparin**<br>**(N=228)** | **Prophylactic**<br>**Heparin**<br>**(N=237)** |
| --- | --- | --- |
| Age (years), mean (SD) | 60.4 (14.1) | 59.6 (15.5) |
| Male sex - no. (%) | 123 (53.9) | 141 (59.5) |
| Race/Ethnicity - no. (%)† |  |  |
| White |  |  |
| European | 97 (43.7) | 96 (40.9) |
| Middle Eastern, North African | 65 (29.3) | 67 (28.5) |
| Asian | 27 (12.2) | 38 (16.2) |
| Black or African American | 18 (8.1) | 23 (9.8) |
| Hispanic or Latino | 14 (6.3) | 10 (4.3) |
| American Indian, Alaska Native, First Nations,<br>Indigenous/Aboriginal, Metis | 0 (0.0) | 1 (0.4) |
| Native Hawaiian or Other Pacific Islander | 1 (0.5) | 0 (0.0) |
| Body mass index (kg/m<sup>2</sup>), mean (SD)‡ | 30.3 (6.4) | 30.2 (7.0) |
| Duration of symptoms prior to hospitalization (days), mean (SD)¶ | 7.1 (5.1) | 7.1 (5.2) |
| Duration of hospitalization before randomization (days), mean (SD) | 1.5 (1.1) | 1.4 (1.0) |
| Hypoxia at baseline - no. (%)<sup>§</sup> | 190 (90.9) | 203 (93.1) |
| Pre-existing conditions - no. (%) |  |  |
| Hypertension | 108 (47.4) | 117 (49.4) |
| Diabetes mellitus | 83 (36.4) | 77 (32.5) |
| Coronary artery disease | 16 (7.0) | 18 (7.6) |
| Heart failure | 9 (3.9) | 6 (2.5) |
| Atrial fibrillation | 0 (0.0) | 2 (0.8) |
| Cerebrovascular disease | 10 (4.4) | 9 (3.8) |
| Peripheral vascular disease | 0 (0.0) | 1 (0.4) |
| Past history of venous thromboembolism | 3 (1.3) | 2 (0.8) |
| Chronic pulmonary disease§ | 36 (15.8) | 27 (11.4) |
| Chronic kidney disease | 20 (8.8) | 13 (5.5) |
| Chronic liver disease | 5 (2.2) | 9 (3.8) |
| Cancer | 13 (5.7) | 19 (8.0) |
| Immunodeficiency | 1 (0.4) | 2 (0.8) |
| Autoimmune disease | 6 (2.6) | 11 (4.6) |
| Cognitive impairment | 12 (5.3) | 11 (4.6) |
| Mental illness | 18 (7.9) | 13 (5.5) |
| Active smoking | 5 (2.2) | 7 (3.0) |
| Medication history - no. (%)** |  |  |
| Systemic corticosteroids | 161 (70.6) | 162 (68.4) |
| Antiplatelet agent | 24 (10.5) | 29 (12.2) |
| Prior COVID-19 vaccine administration - no. (%) | 1 (0.4) | 2 (0.8) |
| Laboratory values |  |  |
| D-dimer |  |  |
| D-dimer positivity - no. (%)*** | 222 (97.4) | 232 (97.9) |
| D-dimer x ULN, geometric mean (SD)<sup>°</sup> | 2.1 (0.7) | 2.5 (0.9) |
| Distribution - no. (%) |  |  |
| D-dimer <2 x ULN | 115 (50.4) | 112 (47.3) |
| D-dimer ≥ 2 - 3 x ULN | 61 (26.8) | 55 (23.2) |
| D-dimer ≥ 3 - 4 x ULN | 25 (11.0) | 27 (11.4) |
| D-dimer ≥ 4 x ULN | 27 (11.8) | 43 (18.1) |
| Platelet count (10<sup>9</sup>/L), mean (SD)<sup>∞</sup> | 233.7 (95.7) | 237.8 (95.3) |
| Creatinine (μmol/L), mean (SD)<sup>°</sup> | 84.6 (44.1) | 85.9 (58.2) |
| Country - no. (%) |  |  |
| Brazil | 54 (23.7) | 51 (21.5) |
| Canada | 72 (31.6) | 78 (32.9) |
| Ireland | 11 (4.8) | 12 (5.1) |
| Saudi Arabia | 71 (31.1) | 76 (32.1) |
| United Arab Emirates | 7 (3.1) | 6 (2.5) |
| United States of America | 13 (5.7) | 14 (5.9) |
| Enrolled in another COVID-19 trial - no. (%) | 29 (12.7) | 31 (13.1) |

\*SD: standard deviation; ULN: upper limit of normal.
†Data regarding race/ethnicity were missing for 6 patients in the therapeutic heparin group and 2 patients for the prophylactic heparin group.
‡Body-mass index (BMI) is the weight in kilograms divided by the square of the height in meters; Data regarding BMI was missing for 6 participants in the therapeutic heparin group and 4 participants in the prophylactic heparin group.
¶Data regarding duration of symptoms prior to hospitalization were missing for 1 patient in the therapeutic heparin group and 5 for the prophylactic heparin group.
δHypoxia was defined as oxygen saturation <93% on room air; Data regarding hypoxia was missing for 19 patients in the therapeutic heparin and 19 patients in the prophylactic heparin.
§Includes chronic restrictive pulmonary disease, chronic obstructive pulmonary disease, and asthma.
\*\*No patients were on remdesivir or tocilizumab at baseline.
\*\*\*6 patients in the therapeutic heparin group and 5 prophylactic heparin group had D-dimer levels below the ULN.
<sup>°</sup>SD for the natural logarithm of D-dimer levels x ULN.
<sup>∞</sup>Data regarding platelet count was missing for 16 patients in the therapeutic heparin group and 24 patients in the prophylactic heparin group.
<sup>°</sup>Data regarding creatinine was missing for 14 patients in the therapeutic heparin group and 23 patients in the prophylactic heparin group.

### Primary and Secondary Outcomes

The primary outcome occurred in 37 patients (16.2%) assigned to therapeutic heparin and 52 patients (21.9%) assigned to prophylactic heparin (odds ratio, 0.69; 95% confidence interval [CI], 0.43 to 1.10, p=0.12; Table 2). Death from any cause occurred in four (1.8%) with therapeutic heparin versus 18 (7.6%) with prophylactic heparin (odds ratio, 0.22; 95%-CI, 0.07 to 0.65). Hypoxic respiratory failure was the most common cause of death (Table S3). The composite of invasive or non-invasive occurred in 21 patients (9.2%) with therapeutic heparin and 26 (11.0%) with prophylactic heparin (odds ratio, 0.82; 95%-CI, 0.45 to 1.51). ICU admission occurred in 33 patients (14.5%) with therapeutic heparin and 42 (17.7%) with prophylactic heparin (odds ratio, 0.79; 95%-CI, 0.48 to 1.29). The composite of all-cause death or invasive/non-invasive mechanical ventilation occurred in 23 (10.1%) patients with therapeutic heparin and 38 (16.0%) with prophylactic heparin (odds ratio, 0.59; 95%-CI, 0.34 to 1.02).

**Table 2.** Primary and Secondary Outcomes*.

| **Outcome** | **Therapeutic Heparin (N=228)**<br>*no. of patients (%)* | **Prophylactic Heparin (N=237)**<br>*no. of patients (%)* | **Odds Ratio or Ratio of Geometric Means<sup>‡</sup> (95%-CI)** |
| --- | --- | --- | --- |
| Primary Composite Outcome<sup>†</sup> | 37 (16.2) | 52 (21.9) | 0.69 (0.43, 1.10) |
| Components of the primary composite outcome |  |  |  |
| Death from any cause | 4 (1.8) | 18 (7.6) | 0.22 (0.07, 0.65) |
| Invasive mechanical ventilation | 11 (4.8) | 16 (6.8) | 0.70 (0.32, 1.55) |
| Any mechanical ventilation<sup>°</sup> | 21 (9.2) | 26 (11.0) | 0.82 (0.45, 1.51) |
| ICU admission | 33 (14.5) | 42 (17.7) | 0.79 (0.48, 1.29) |
| Death or any mechanical ventilation | 23 (10.1) | 38 (16.0) | 0.59 (0.34, 1.02) |
| Death or ICU admission | 36 (15.8) | 50 (21.1) | 0.70 (0.44, 1.13) |
| Ventilator-free days alive (days), mean (SD) | 26.5 (5.6) | 24.7 (8.5) | 1.77 (1.02, 3.08) |
| Organ support-free days alive (days), mean (SD) | 25.8 (6.2) | 24.1 (8.8) | 1.41 (0.90, 2.21) |
| ICU-free days alive (days), mean (SD) | 26.0 (6.1) | 24.2 (8.8) | 1.51 (0.94, 2.41) |
| Hospital-free days alive (days), mean (SD) | 19.8 (7.3) | 18.4 (9.2) | 1.09 (0.79, 1.5) |
| Renal replacement therapy<sup>¶</sup> | 2 (0.9) | 5 (2.1) | 0.41 (0.08, 2.15) |
| Thromboembolism<sup>*</sup> |  |  |  |
| Venous | 2 (0.9) | 7 (3.0) | 0.29 (0.06, 1.42) |
| Arterial | 0 (0.0) | 1 (0.4) | - |
| Bleeding |  |  |  |
| ISTH major bleeding<sup>§</sup> | 2 (0.9) | 4 (1.7) | 0.52 (0.09, 2.85) |
| Red blood cell transfusion ($\geq 1$ unit) | 3 (1.3) | 9 (3.8) | 0.34 (0.09, 1.27) |
| Transfusion of hemostatic blood components or products<sup>‡</sup> | 1 (0.4) | 0 (0.0) | - |
| Heparin induced thrombocytopenia | 0 (0.0) | 0 (0.0) | - |
| D-dimer x ULN, geometric mean (SD)<sup>‡</sup> | 1.9 (0.7) | 2.4 (0.9) | 0.88 (0.78, 0.99) |

\*ICU: intensive care unit; CI: confidence interval; SD: standard deviation; ISTH: International Society on Thrombosis and Haemostasis.
All outcomes were assessed up to 28 days post-randomization.
<sup>†</sup>Defined as death, invasive mechanical ventilation, non-invasive mechanical ventilation or ICU admission.
<sup>°</sup>Invasive or non-invasive (bilevel or continuous positive airway pressure) mechanical ventilation.
<sup>‡</sup>Transfusion of platelets, frozen plasma, prothrombin complex concentrate, cryoprecipitate and/or fibrinogen concentrate; 17 patients received convalescent plasma and were not included in the count.
<sup>§</sup>Major bleeding defined by the ISTH Scientific and Standardization Committee.
<sup>¶</sup>Continuous renal replacement therapy or intermittent hemodialysis.
<sup>\*</sup>All diagnostically confirmed except for 1 symptomatic deep vein thrombosis in the prophylactic heparin group, which could not be definitively confirmed as diagnostic imaging was not done during acute symptomatic period.
<sup>‡</sup>Ratio of geometric means of D-dimer levels x ULN of day 2 ± 24h post-randomization, adjusted for baseline geometric means of D-dimer levels x ULN using analysis of covariance. SD for the natural logarithm of D-dimer levels x ULN. The day 2 ± 24 hours D-dimer was missing for 66 in the therapeutic heparin group and 64 in the prophylactic heparin group.

The mean number of ventilator-free days was 26.5 (standard deviation, 5.6) with therapeutic heparin, and 24.7 (standard deviation, 8.5) with prophylactic heparin (odds ratio from ordinal logistic regression, 1.77; 95%-CI, 1.02 to 3.08). The mean number of organ support-free days was 25.8 (standard deviation, 6.2) with therapeutic heparin, and 24.1 (standard deviation, 8.8) with prophylactic heparin (odds ratio, 1.41; 95%-CI, 0.90 to 2.21). The mean number of ICU-free days was 26.0 (standard deviation, 6.1) with therapeutic heparin, and 24.2 (standard deviation, 8.8) with prophylactic heparin (odds ratio, 1.51; 95%-CI, 0.94 to 2.41). There was no relevant between-group difference in hospital-free days (Table 2).

The number of venous thromboembolic events was 2 (0.9%) with therapeutic heparin and 7 (3.0%) with prophylactic heparin (odds ratio, 0.29, 95%-CI, 0.06 to 1.42; Tables 2 and S4). There were no fatal thromboembolic events. D-dimer levels, assessed at a median of 1.5 days (interquartile range, 1 to 2) in 162 (71.1%) and 173 patients (73.0%) assigned to therapeutic and prophylactic heparin, respectively, were lower with therapeutic heparin (geometric mean ratio, 0.88; 95%-CI, 0.78 to 0.99).

Subgroup analyses of the primary outcome did not provide evidence for differences in treatment effect between subgroups (Fig. S2). Findings from the per-protocol analyses and other sensitivity analyses were similar to those from the primary analysis (Table S5 to S12).

### Safety Outcomes

There were 2 patients with major bleeding events in the therapeutic heparin group and 4 in the prophylactic heparin group (odds ratio 0.52; 95%-CI, 0.09 to 2.85; Table 2). There were no fatal bleeding events and there were no cases of intracranial hemorrhage (Tables S13 and S14).

### Meta-analyses

There were 3 trials of therapeutic heparin compared to usual care using lower doses of heparin. Two trials included moderately ill ward patients with Covid-19, the RAPID trial and a multiplatform trial integrating the Antithrombotic Therapy to Ameliorate Complications of Covid-19 (ATTACC), Accelerating Covid-19 Therapeutic Interventions and Vaccines-4 Antithrombotics Inpatient Platform Trial (ACTIV-4a) and the Randomized, Embedded, Multifactorial Adaptive Platform Trial for Community-Acquired Pneumonia (REMAP-CAP).^19^ A separate multiplatform trial conducted by the same investigators evaluated therapeutic heparin in severely ill ICU patients.^20^ Figure 2 shows the meta-analyses of the two trials of moderately ill patients. There was no significant reduction in all-cause death (odds ratio, 0.74; 95%-CI, 0.54 to 1.02), but significant reductions in the composite of death or invasive mechanical ventilation (odds ratio, 0.77; 95%-CI, 0.60 to 0.99), death or organ support (odds ratio, 0.77; 95%-CI, 0.63 to 0.93), death or major thrombotic event (odds ratio, 0.64; 95%-CI, 0.48 to 0.86), and major thrombotic events (odds ratio, 0.47; 95%-CI, 0.25 to 0.87). Ventilator-free days alive (odds ratio, 1.30; 95%-CI 1.05 to 1.61) and organ support-free days alive (odds ratio 1.31, 95% 1.08 to 1.60) were significantly increased with therapeutic heparin. Conversely, there was a non-significant increase in major bleeding. There were significant treatment-by-subgroup interactions with severity of illness (moderately ill ward patients versus severely ill ICU patients) for all-cause death (p for interaction=0.045), all-cause death or major thrombotic event (p=0.018) and organ-support-free days alive (p=0.006; Fig. S3). There was no evidence for treatment-by-subgroup interactions for major thrombotic events and major bleeding.

**Figure 2.**
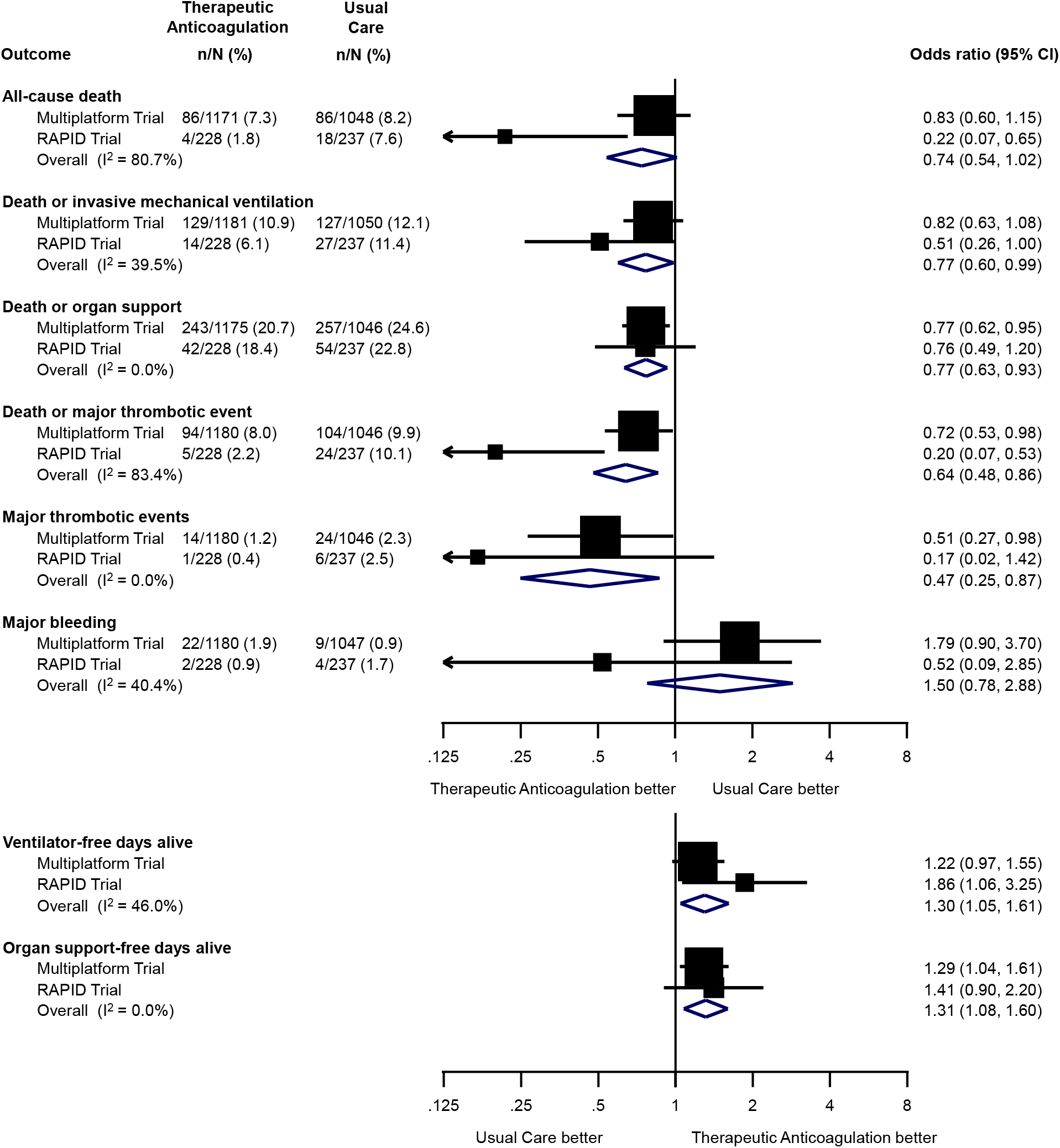
Meta-Analyses of Effectiveness and Safety Outcomes in Randomized Trials Comparing Therapeutic Heparin with Usual Care in Moderately Ill Ward Patients with Covid-19. Mantel-Haenszel fixed-effect meta-analyses of the RAPID trial and the multiplatform trial in moderately ill ward patients.^19^ Squares and horizontal lines show treatment effects and their 95% confidence intervals in each trial. The area of each square is proportional to the weight the trial received in the meta-analysis. Diamonds show estimated treatment effects and 95% confidence intervals from meta-analyses. Odds ratios for ventilator-free and organ support-free days alive are from ordinal logistic regression in both trials; death was assigned the worst outcome (a value of −1). Major thrombotic events were defined as the composite of myocardial infarction, pulmonary embolism, ischemic stroke or systemic arterial embolism; Major bleeding defined by the ISTH Scientific and Standardization Committee. In accordance with the primary outcome definition of the multiplatform trials,^19,20^organ support-free days alive were calculated for an observation time of 21 days; remaining outcomes were based on an observation time of 28 days.

## DISCUSSION

In this randomized trial of moderately ill hospitalized ward patients with Covid-19 and elevated D-dimer levels, therapeutic heparin did not lower the incidence of the primary composite of death, mechanical ventilation or ICU admission compared with prophylactic heparin. The odds of all-cause death were significantly reduced by 78% with therapeutic heparin, however. Between-group differences were smaller and nonsignificant for mechanical ventilation and ICU admission.

A meta-analysis of the two available trials of therapeutic heparin in moderately ill ward patients with Covid-19 did not provide conclusive evidence for a reduction in mortality with therapeutic heparin, with a larger mortality reduction in RAPID than in the multiplatform trial.^19^ Findings for remaining effectiveness outcomes were consistent between trials, with significant differences in favour of therapeutic heparin. Three effectiveness outcomes were also reported in a separate multiplatform trial in severely ill patients.^20^ We found significant treatment-by-subgroup interactions with severity of illness for all-cause death, all-cause death or major thrombosis and organ-support-free days alive, with evidence of benefit with therapeutic heparin in moderately ill ward patients, but not in severely ill ICU patients. Conversely, there were no treatment by subgroup interactions for major thrombotic events and major bleeding, with consistent benefit of therapeutic heparin for major thrombotic events, and non-significant small increases in major bleeding irrespective of illness severity. Both, the multiplatform trial and RAPID used D-dimer levels for risk stratification of ward patients,^21,26^ but results of the multiplatform trial indicate that D-dimer status does not result in variation of treatment effects.^19^ Taken together, these trials provide conclusive evidence for benefit of therapeutic heparin in moderately ill ward patients with Covid-19 irrespective of D-dimer levels, but not in severely ill ICU patients.

The mortality reduction was more pronounced in RAPID than in the multiplatform trial.^19^ Potential explanations include chance and a stronger contrast between treatment arms in RAPID as compared with the multiplatform trial:^19^ for pragmatic reasons, the multiplatform trial allowed intermediate dose heparin in the control group, and 28.2% received higher than prophylactic doses.^19^ RAPID only allowed prophylactic heparin doses, and only 2.1% received higher doses (Table S15). The effectiveness of anticoagulation also seems to depend on the type of anticoagulant: the Anticoagulation Coronavirus (ACTION) trial used 15 to 20 mg of rivaroxaban in 94% of patients assigned to therapeutic anticoagulation and found no benefit.^27^ Rivaroxaban is unlikely to have the anti-inflammatory and anti-viral properties attributed to heparin.^2,11–15^ In addition, ACTION allowed intermediate doses of enoxaparin in the control group.

Our trial has two major limitations. First, RAPID had an adaptive design. The protocol prespecified that the sample size would be increased if the conditional power at 75% of the original sample size was between 60 and 80%.^21^ However, the conditional power was below 60%, therefore the sample size was not increased, thus RAPID remained underpowered. Second, the trial had an open-label design, but all relevant outcomes were blindly adjudicated by an independent clinical events committee.

The RAPID trial did not find a significant reduction in the primary composite outcome of death, mechanical ventilation or ICU admission with therapeutic heparin. However, in conjunction with the multiplatform trial,^19^ it suggests a clinical benefit of therapeutic heparin in moderately ill ward patients with Covid-19.

### Support

Funded by Task 54, Defence Research Development Canada, Department of National Defence, Ottawa, Canada; St. Michael’s Hospital Foundation, Toronto, Canada; St. Joseph’s Health Centre Foundation, Toronto, Canada; 2020 TD Community Health Solutions Fund – COVID-19 Research Grant; Michael Garron Hospital, Toronto, Canada; The Ottawa Hospital Foundation COVID-19 Emergency Response Fund, Ottawa, Canada; International Network of Venous Thromboembolism Clinical Research Networks (INVENT) Kickstarter Award; Science Foundation Ireland, Enterprise Ireland, IDA Ireland COVID-19 Rapid Response Funding Call 20/COV/0157; SEAMO (Southeastern Ontario Academic Medical Organization) COVID-19 Innovation Fund; P20 GM135007 from the National Institute of General Medical Sciences, NIH; University of Vermont Medical Center Fund Grant; College of Medicine Research Center, Deanship of Scientific Research, King Saud University, Riyadh, Saudi Arabia.

## Data Availability

Data may be available upon request in accordance with appropriate privacy and data sharing principles.

## Acknowledgements

We are grateful to all patients and hospital staff who contributed to this trial. We are indebted to the individual donors who contributed to RAPID crowd funding; hospital sites that donated research infrastructure in kind, and the data and safety monitoring board.

## References

1. Richardson S, Hirsch JS, Narasimhan M, Crawford JM, McGinn T, Davidson KW, et al. Presenting Characteristics, Comorbidities, and Outcomes Among 5700 Patients Hospitalized With COVID-19 in the New York City Area. JAMA. 2020 May 26;323(20):2052–9.

2. Tang N, Bai H, Chen X, Gong J, Li D, Sun Z. Anticoagulant treatment is associated with decreased mortality in severe coronavirus disease 2019 patients with coagulopathy. J Thromb Haemost. 18(5):1094–9.

3. Guan W, Ni Z, Hu Y, Liang W, Ou C, He J, et al. Clinical Characteristics of Coronavirus Disease 2019 in China. N Engl J Med. 2020 Feb 28;NEJMoa2002032.

4. McGonagle D, O’Donnell JS, Sharif K, Emery P, Bridgewood C. Immune mechanisms of pulmonary intravascular coagulopathy in COVID-19 pneumonia. Lancet Rheumatol. 2020 Jul;2(7):e437–45.

5. Wichmann D, Sperhake J-P, Lütgehetmann M, Steurer S, Edler C, Heinemann A, et al. Autopsy Findings and Venous Thromboembolism in Patients With COVID-19: A Prospective Cohort Study. Ann Intern Med. 2020 May 6;

6. Negri EM, Piloto BM, Morinaga LK, Jardim CVP, Lamy SAE-D, Ferreira MA, et al. Heparin Therapy Improving Hypoxia in COVID-19 Patients - A Case Series. Front Physiol. 2020;11:573044.

7. Tang N, Li D, Wang X, Sun Z. Abnormal coagulation parameters are associated with poor prognosis in patients with novel coronavirus pneumonia. J Thromb Haemost. 2020 Apr;18(4):844–7.

8. Petrilli CM, Jones SA, Yang J, Rajagopalan H, O’Donnell L, Chernyak Y, et al. Factors associated with hospital admission and critical illness among 5279 people with coronavirus disease 2019 in New York City: prospective cohort study. BMJ. 2020 May 22;369:m1966.

9. Thachil J, Cushman M, Srivastava A. A Proposal for Staging COVID-19 Coagulopathy. Research and Practice in Thrombosis and Haemostasis. 4(5):731–6.

10. Iba T, Levy JH, Levi M, Connors JM, Thachil J. Coagulopathy of Coronavirus Disease 2019. Crit Care Med. 2020 May 26;48(9):1358–64.

11. Mu S, Liu Y, Jiang J, Ding R, Li X, Li X, et al. Unfractionated heparin ameliorates pulmonary microvascular endothelial barrier dysfunction via microtubule stabilization in acute lung injury. Respir Res. 2018 Nov 15;19(1):220.

12. Li L, Ma X, Li X. [Effect of heparin on histone-mediated the expression of von Willebrand factor and fibrinogen in lung tissue]. Zhonghua Wei Zhong Bing Ji Jiu Yi Xue. 2019 Nov;31(11):1363–7.

13. Liu J, Li J, Arnold K, Pawlinski R, Key NS. Using heparin molecules to manage COVID-2019. Research and Practice in Thrombosis and Haemostasis. 4(4):518–23.

14. Helms J, Tacquard C, Severac F, Leonard-Lorant I, Ohana M, Delabranche X, et al. High risk of thrombosis in patients with severe SARS-CoV-2 infection: a multicenter prospective cohort study. Intensive Care Med. 2020 May 4;

15. Wang C, Chi C, Guo L, Wang X, Guo L, Sun J, et al. Heparin therapy reduces 28-day mortality in adult severe sepsis patients: a systematic review and meta-analysis. Crit Care. 2014 Oct 16;18(5):563.

16. Zhou F, Yu T, Du R, Fan G, Liu Y, Liu Z, et al. Clinical course and risk factors for mortality of adult inpatients with COVID-19 in Wuhan, China: a retrospective cohort study. The Lancet. 2020 Mar;395(10229):1054–62.

17. Nopp S, Moik F, Jilma B, Pabinger I, Ay C. Risk of venous thromboembolism in patients with COVID-19: A systematic review and meta-analysis. Res Pract Thromb Haemost. 2020 Sep 25;4(7):1178–91.

18. Paranjpe I, Fuster V, Lala A, Russak AJ, Glicksberg BS, Levin MA, et al. Association of Treatment Dose Anticoagulation With In-Hospital Survival Among Hospitalized Patients With COVID-19. J Am Coll Cardiol. 2020 Jul 7;76(1):122–4.

19. Lawler PR, Goligher EC, Berger JS, Neal MD, McVerry BJ, Nicolau JC, et al. Therapeutic Anticoagulation in Non-Critically Ill Patients with Covid-19. medRxiv. 2021 Jan 1;2021.05.13.21256846.

20. Goligher E, Bradbury C, McVerry BJ, Lawler PR, Berger J, Gong MN, et al. Therapeutic Anticoagulation in Critically Ill Patients with Covid-19 – Preliminary Report. medRxiv. 2021 Jan 1;2021.03.10.21252749.

21. Sholzberg M, Tang GH, Negri E, Rahhal H, Kreuziger LB, Pompilio CE, et al. Coagulopathy of hospitalised COVID-19: A Pragmatic Randomised Controlled Trial of Therapeutic Anticoagulation versus Standard Care as a Rapid Response to the COVID-19 Pandemic (RAPID COVID COAG – RAPID Trial): A structured summary of a study protocol for a randomised controlled trial. Trials. 2021 Mar 10;22(1):202.

22. Schulman S, Kearon C. Definition of major bleeding in clinical investigations of antihemostatic medicinal products in non-surgical patients. Journal of Thrombosis and Haemostasis. 2005;3(4):692–4.

23. Doi SA, Furuya-Kanamori L, Xu C, Lin L, Chivese T, Thalib L. Questionable utility of the relative risk in clinical research: a call for change to practice. J Clin Epidemiol. 2020 Nov 7;

24. REMAP-CAP Investigators, Gordon AC, Mouncey PR, Al-Beidh F, Rowan KM, Nichol AD, et al. Interleukin-6 Receptor Antagonists in Critically Ill Patients with Covid-19. N Engl J Med. 2021 Apr 22;384(16):1491–502.

25. WHO Solidarity Trial Consortium, Pan H, Peto R, Henao-Restrepo A-M, Preziosi M-P, Sathiyamoorthy V, et al. Repurposed Antiviral Drugs for Covid-19 - Interim WHO Solidarity Trial Results. N Engl J Med. 2021 Feb 11;384(6):497–511.

26. Houston BL, Lawler PR, Goligher EC, Farkouh ME, Bradbury C, Carrier M, et al. Anti-Thrombotic Therapy to Ameliorate Complications of COVID-19 (ATTACC): Study design and methodology for an international, adaptive Bayesian randomized controlled trial. Clin Trials. 2020 Oct;17(5):491–500.

27. Lopes RD, de Barros E Silva PGM, Furtado RHM, Macedo AVS, Bronhara B, Damiani LP, et al. Therapeutic versus prophylactic anticoagulation for patients admitted to hospital with COVID-19 and elevated D-dimer concentration (ACTION): an open-label, multicentre, randomised, controlled trial. Lancet. 2021 Jun 12;397(10291):2253–63.

