## Supplementary Appendix for "Heparin for Moderately Ill Patients with Covid-19"

This appendix has been provided by the authors to give readers additional information about their work.

**Data and Safety Monitoring Board**: Andreas Laupacis (Chair), Yulia Lin, Andrew Day, Bram Rochwerg

**Medical Monitors:** Eric K. Tseng, Gloria Lim

**Independent Event-Adjudication Committee:** Jameel Abdulrehman (English), Mansour Gergi (English), Rodrigo Hidd Kondo (Portuguese), Bruna Mamprim Piloto (Portuguese)

**Study Investigators and Coordinators:**

**Brazil:**
*Hospital das Clinicas HCFMUSP, Faculdade de Medicina, Universidade de Sao Paulo, SP, BR*: Elnara Márcia Negri, Hassan Rahhal, Carlos Eduardo Pompílio, Guilherme de Abreu Pereira, Fernando Salvetti Valente, Ariel Fernando Villarroel Agreda, Pablo Andres Munoz Torres, Maíra Oliveira Moraes, Claudia de Lucena Moreira, Fernando Galassi Stocco Neto, Fabíola Vieira Duarte Baptista, Joanne Alves Moreira, Augusto Séttemo Ferreira, Paula Frudit, Gabriel Martinez, Heraldo Possolo Souza, Rodrigo Antônio Brandão Neto, Elbio Antonio d'Amico, Eduardo Messias Hirano Padrão, Fernando Sarin da Mota e Albuquerque, Giovanna Villela Zangrossi, Alberto Kendy Kanasiro, Alissom Vitti Cincoto, Hugo de Souza Reis, Vitor Miyashiro Arias da Silva, Arthur Petrillo Bellintani, Vivian dos Santos Pereira, Yohan Washington de Oliveira, Bárbara Justo Carvalho, Mariana de Souza Novaes, Henrique Brito Silveira, Gabriel César Alves de Avelar, Lara Bonanni Mota, Karina Caciola, Matheus de Arêa Leão Freire Marim, Tales Cabral Monsalvarga, Alexandre Salgado Blanco Santos, Ahmed Haydar, Gabriella Seixas Sampaio Saraiva, Sabrina Correa da Costa Ribeiro, Julio Flavio Meirelles Marchini, Julio Cesar Garcia de Alencar, Ana Catharina de Seixas Santos

**Canada:**
*Hôpital Charles Lemoyne, Longueuil, QC:* Catherine Sperlich, Caroline Millette, Mathieu Lebeau, Céline Devaux, Mélina Boutin, Trung Nghia Nguyen, Line Srour, Flavia de Angelis, Mariane Fugulin

*William Osler Health System (Brampton Civic Hospital and Etobicoke General Hospital), Brampton and Etobicoke, ON:* Sabrena Tangri, Alexandra Binnie, Shayna A.D. Bejaimal, Andrew Binding, Rosa M. Marticorena, Galo Ginocchio

*Mount Sinai Hospital, Toronto, ON:* Michael Fralick, Eric Kaplovitch, Klaudia Rymaszewski, Afsaneh Raissi, Marcelo Falappa

*Trillium Health Partners (Credit Valley Hospital and Mississauga Hospital)*, Mississauga, ON: Terence Tang, Blair Ernst, Amna Ali, Martin Romano, Mobina Khurram

*St. Joseph's Health Centre, Toronto, ON:* Peter Jaksa, Christie Kim, Ajay Kapur, Michelle Edwards, Vidushi Swarup, Bruna Camilotti, Jiten Jani, Jeff Carter

*Alberta Health Services (Foothills Medical Centre, Rockyview General Hospital, Peter Lougheed Centre), Calgary, AB:* Deepa Suryanarayan, Mark R. Gillrie, Davinder Sidhu, Traci Robinson

*The Ottawa Hospital (General and Civic Campus), Ottawa, ON:* Lana Castellucci, Krystina Stutely, Irene Watpool, Rebecca Porteous

*Hôpital du Sacré-Coeur de Montréal, Montréal, QC*: Karine Doyon, Kevin Jao, Jean-Samuel Boudreault-Pedneault

*Michael Garron Hospital, Toronto, ON:* E. Roseann Andreou, Christopher Kandel, Maureen T. Taylor, Wei En Enoch Choo

*St. Michael's Hospital, Toronto, ON:* Vera Dounaevskaia, Eric K. Tseng, Gloria Lim, Vidushi Swarup, Laura Parsons, Ann Dowbenka, Gitana Ramonas

*University of Alberta Hospital, Edmonton, AB:* Cynthia Wu, Sergey Nikitin, Jeffery Patterson, Mark Hnatiuk, Anca Tapardel, Sarah Takach- Lapner, Thirza Carpenter, Lori Rackel, Rebecca Cairns, Jessica Pinder, Sergey Nikitin

*Southlake Regional Health Centre, Newmarket, ON:* Jai Jayakar, Peter Anglin, Catherine McPherson, Liselle Chiverton, David Barbosa, Shany Loukiantchenko, Yana Shamiss

*Hospital Maisonneuve-Rosemont, Montréal, QC:* Marie-Pier Arsenault, Danaë Tassy

*Nova Scotia Health Authority (Queen Elizabeth II Health Sciences Centre), Halifax, NS:* Sudeep Shivakumar, Mary-Margaret Keating, Sue Pleasance

**Ireland:**
*Mater Misericordiae University Hospital, Dublin and University College Dublin Clinical Research Centre:* Fionnuala Ní Áinle, Barry Kevane, Sarah Cullivan, Nick Power, Peter Doran, Kelly Leamy, Aoife Kelly, Conor Moran, Mairead O'Connor, Aoife McDonnell, Roseanne Boyce, Faiza Sefroun, Rabia Hussain, Patrick Murray, Anna Malara, Brenda Molloy, Meadbh O'Halloran, Emer Cunningham, Jack Lambert, Aoife Cotter, Brian Marsh, Gerard Sheehan, Eavan Muldoon, James Woo, Sean Gaine, Deirdre Morley

**Saudi Arabia:**
*King Saud University Medical City, Riyadh*: Musaad AlHamzah, Khalid Alayed, Farjah H AlGahtani, Ibrahim Almaghlouth, Sondus Ata, Fai Alkhathlan, Najma Khalil, Israa Mohamed Hussein, Mohammed Bashir, Ahmed S. BaHammam, Abdulrahman Alsultan, Hadeel Alkofide, Tariq M Alhawassi

*King Fahad Medical City, Riyadh*: Mohammed Alsheef, Fahad AlSumait, Abdulhadi M. Alqahtani, Emad K. Zayed, Ammar AlSughayir, Yacoub Abuzied

*King Faisal Specialist Hospital, Riyadh*: Faris Alomran, Hazzaa AlZahrani, Jawaher Al-Otaibi, Haya Alothaimeen, Noura Alzannan

**United States of America:**
*Versiti, Milwaukee, Wisconsin*: Lisa Baumann Kreuziger, Amer Al Homssi, Haisam Abid, Stephanie Jones, Shannon Broaddrick, Neha Jain

*University of Vermont Medical Center, Burlington, Vermont:* Christos Colovos, Mary Cushman, Roz King, Mohit Jindal

*Barnes Jewish Hospital, St. Louis, Missouri:* Kristen Sanfilippo, Patty Nieters

**United Arab Emirates:**
*Al Ain Hospital, Al Ain:* Mozah Almarshoodi, Muhammad Hammad, Aleeswa Francis Benny, Tariq A. Hamdan, Suhaib Kamal Ahmed Elobaid, Ibrahim Khafagi, Saima Saeed Ahmed, AbduleRehman AlEssaie, Shamma AlAlawi, Khloud Bashir, Aysha Abdulla Salem Al Suwaidi, Hiba Ibrahim Khogali Ahmed, Mohamed Milad Ismail, El Mutasim Ahmed El Faki

**Data Management and Coordination Center (DMCC):**
*Applied Health Research Centre, St. Michael's Hospital/University of Toronto:* Mercy Charles, Alice Dang, Gurpreet Lakhanpal, Dominic Lee, Prachi Ray, Maria Naydenova, Kosma Wysocki

*Hematology-Oncology Research Group, St. Michael's Hospital/University of Toronto:* Michelle Sholzberg, Aziz Jiwajee, Vidushi Swarup, Bruna Camilotti, Grace H. Tang

**Regional Coordination Centers:**
*US Coordination Center (Versiti, Milwaukee, Wisconsin):* Lisa Baumann Kreuziger, Stephanie Jones, Shannon Broaddrick, Greg Wendling

*Saudi Arabia Coordination Center (King Saud University Medical City, Riyadh):* Musaad AlHamzah, Ahmed S. BaHammam, Abdulrahman Alsultan, Hadeel Alkofide, Tariq M Alhawassi

*European Legal Representative* (*Mater Misericordiae University Hospital, Dublin):* Fionnuala Ní Áinle, Brenda Molloy, Anna Malara, Emer Cunningham, Meadbh O'Halloran, Aoife Cotter, Brian Marsh, Jack Lambert, Gerard Sheehan, Eavan Muldoon, James Woo, Sean Gaine

*Brazil Coordination Center (Hospital das Clínicas da Faculdade de Medicina da Universidade de São Paulo, São Paulo):* Hassan Rahhal, Elnara Márcia Negri, Carlos Eduardo Pompilio, Fernando S. Valente and Guilherme de Abreu Pereira.

**Statistical Analysis (AHRC)**: Bruno R da Costa, Kevin E. Thorpe, Fei Zuo, Peter Jüni

**Biorepository Coordination Center:**
*Laboratory for Clinical Biochemistry Research, University of Vermont Larner College of Medicine, Burlington, Vermont:* Mary Cushman, Rebekah Boyle, Elaine Cornell, Debora Kamin Mukaz

### 2. Funding Agencies

**The RAPID Trial was funded by:**

- Task 54, Defence Research Development Canada, Department of National Defence, Ottawa, Canada
- St. Michael's Hospital Foundation, Toronto, Canada
- St. Joseph's Health Centre Foundation, Toronto, Canada
- 2020 TD Community Health Solutions Fund – COVID-19 Research Grant, Michael Garron Hospital, Toronto, Canada
- The Ottawa Hospital Foundation COVID-19 Emergency Response Fund, Ottawa, Canada
- International Network of Venous Thromboembolism Clinical Research Networks (INVENT) Kickstarter Award
- Science Foundation Ireland, Enterprise Ireland, IDA Ireland COVID-19 Rapid Response Funding Call 20/COV/0157
- Southeastern Ontario Academic Medical Organization (SEAMO) COVID-19 Innovation Fund
- P20 GM135007 from the National Institute of General Medical Sciences, NIH
- University of Vermont Medical Center Fund Grant
- College of Medicine Research Center, Deanship of Scientific Research, King Saud University, Riyadh, Saudi Arabia

**Research Personnel and Infrastructure in kind provided by:**

- Sinai Health Foundation, Sinai Health, Toronto, Ontario
- Trillium Health Partners Pharmacy Team and Institute for Better Health, Trillium Health Partners, Toronto, Ontario
- University College Dublin Clinical Research Centre, Dublin, Ireland
- Nursing and Medical Staff, Patients of Mater Misericordiae University Hospital, Dublin, Ireland
- Versiti Blood Research Institute, Milwaukee, Wisconsin
- Division of Hematology research fund, University of Calgary
- University of Calgary Clinical Research Fund
- Calgary Health Trust Fund
- CMO office Dr. Ghazala Belal Balhaj, AlAin Hospital, AlAin, UAE
- Dr. Ghanem AlHassani, Research committee at SEHA Institute, Abu Dhabi, UAE
- Research committee at the Department of Health (DOH), Abu Dhabi, UAE
- Shawn Rhind PhD and Henry Peng PhD, Defence Research Development Canada, Toronto, Canada
- College of Medicine Research Center, King Saud University, Riyadh, Saudi Arabia
- Clinical Trials Unit, King Saud University Medical City, Riyadh, Saudi Arabia

### 3. Supplementary Methods

#### 3.1 Eligibility Criteria

The inclusion criteria:

1) Laboratory confirmed COVID-19 (diagnosis of SARS-CoV-2 via reverse transcriptase polymerase chain reaction as per the World Health Organization protocol or by nucleic acid based isothermal amplification). Positive test prior to hospital admission **OR** within first 5 days (i.e. 120 hours) after hospital admission;
2) Admitted to hospital for COVID-19;
3) One D-dimer value above ULN (within 5 days (i.e. 120 hours) of hospital admission) **AND EITHER**:
    a. D-Dimer ≥2 times ULN **OR**
    b. D-Dimer above ULN and Oxygen saturation ≤ 93% on room air;
4) ≥ 18 years of age;
5) Informed consent from the patient (or legally authorized substitute decision maker).

The exclusion criteria:

1) pregnancy;
2) hemoglobin <80 g/L in the last 72 hours;
3) platelet count <50 x $10^9$/L in the last 72 hours;
4) known fibrinogen <1.5 g/L (if testing deemed clinically indicated by the treating physician prior to the initiation of anticoagulation);
5) known INR >1.8 (if testing deemed clinically indicated by the treating physician prior to the initiation of anticoagulation);
6) patient already on intermediate dosing of LMWH that cannot be changed (determination of what constitutes an intermediate dose is to be at the discretion of the treating clinician taking the local institutional thromboprophylaxis protocol for high risk patients into consideration);
7) patient already on therapeutic anticoagulation at the time of screening (low or high dose nomogram UFH, LMWH, warfarin, direct oral anticoagulant (any dose of dabigatran, apixaban, rivaroxaban, edoxaban);
8) patient on dual antiplatelet therapy, when one of the agents cannot be stopped safely;
9) known bleeding within the last 30 days requiring emergency room presentation or hospitalization;
10) known history of a bleeding disorder of an inherited or active acquired bleeding disorder;
11) known history of heparin-induced thrombocytopenia;
12) known allergy to UFH or LMWH;
13) admitted to the intensive care unit at the time of screening;
14) treated with non-invasive positive pressure ventilation or invasive mechanical ventilation at the time of screening (of note: high flow oxygen delivery via nasal cannula is acceptable and is not an exclusion criterion);
15) Imminent death according to the judgement of the most responsible physician;
16) enrollment in another clinical trial of antithrombotic therapy involving pre-intensive care unit hospitalized patients.

#### 3.2 Description of Therapeutic Heparin vs. Prophylactic Heparin

Therapeutic Heparin

Therapeutic anticoagulation with low molecular weight heparin (LMWH) or unfractionated heparin (UFH). The choice of LMWH versus UFH was at the clinician's discretion and dependent on local institutional supply. LMWH options are described in Table 1 . UFH was administered using a weight-based nomogram (bolus plus continuous infusion) with activated partial thromboplastin time (aPTT) or UFH anti-Xa titration according to the center-specific institutional protocols as per venous thromboembolism treatment (i.e. high dose nomogram). UFH anti-Xa titration was preferred over aPTT if available as achieving a therapeutic aPTT may be challenging in patients with a pro-inflammatory state such as COVID-19. Therapeutic heparin was administered until hospital discharge, death, day 28 or study withdrawal. If the patient was admitted to the intensive care unit (ICU) or required mechanical ventilatory support (i.e. patient reached a component of the primary composite outcome), continuation of the allocated treatment was recommended, as long as the treating physician was in agreement.

**Table 1. Therapeutic Heparin**
Any of the following strategies could have been used for therapeutic anticoagulation:

| CrCl | BMI | Enoxaparin | Dalteparin | Tinzaparin | UFH |
|---|---|---|---|---|---|
| ≥30 | <40 | 1 mg/kg SC q12h OR<br>1.5 SC mg/kg q24h | 200 units/kg SC q24h OR<br>100 IU/kg SC q12h | 175 U/kg SC q24h | IV bolus, with continuous infusion to titrate to institution specific anti-Xa or aPTT values* |
| | ≥40 | 1 mg/kg q12h[&] | 100 units/kg SC q12h[&] | 175 U/kg SC daily[&] | |
| <30 | <40 | UFH IV bolus, with continuous infusion to titrate to institution specific anti-Xa or aPTT values* or LMWH per hospital protocol taking BMI into consideration as above | | | |
| | ≥40 | | | | |

Abbreviations: CrCl = creatinine clearance; BMI = body mass index; * Initial bolus dose determined by sites, encouraging use of dosing algorithm designed for treatment of venous thromboembolism. UFH anti-Xa titration was preferred over aPTT if available as achieving a therapeutic aPTT may be challenging in patients with a pro-inflammatory state such as COVID-19
[&]For patients with BMI above 40, measurement of anti-Xa to confirm therapeutic effect was suggested.

Prophylactic Heparin

Administration of LMWH, UFH or fondaparinux at thromboprophylactic doses for acutely ill hospitalized medical patients, in the absence of contraindication, is generally considered standard care. The doses of thromboprophylaxis only included those listed below (Table 2).

Any of the following strategies could be used for prophylactic heparin doses above those listed was not considered as prophylactic. A lower dose of either LMWH or UFH than listed below was considered acceptable if due to extremely low weight/BMI, and considered as part of prophylactic heparin:

**Table 2. Prophylactic Heparin**

| CrCl | BMI | Enoxaparin | Dalteparin | Tinzaparin | Fondaparinux | Unfractionated Heparin (UFH) |
|---|---|---|---|---|---|---|
| ≥30 | <40 | 40 mg SC q24h | 5000 units SC q24h | 4500 U SC q24h | 2.5 mg SC q24h | 5000 units SC q8-12h |
| | ≥40 | 40 mg SC q12h | 5000 units SC q12h | 9000 (+/- 1000) U SC q24h | not applicable | 7500 units SC q8h |
| <30 | <40 | UFH 5000 units SC q8-12h or LMWH per hospital protocol taking BMI into consideration as above | | | | |
| | ≥40 | UFH 7500 units SC q8h or LMWH per hospital protocol taking BMI into consideration as above | | | | |

Abbreviations: CrCl = creatinine clearance; BMI = body mass index

Full therapeutic dose anticoagulation (therapeutic dose UFH or LMWH) was permitted as rescue therapy in the event of suspected or confirmed thromboembolism.

**3.3 Primary Outcome**

**Pre-specified primary composite outcome:** ICU admission, non-invasive positive pressure ventilation, invasive mechanical ventilation or death at 28 days.

If a patient was discharged alive before 28 days, vital status was determined using a telephone follow-up. If a patient was discharged alive on mechanical ventilation (invasive or non-invasive) prior to day 28, a call to the patient or a doctor/nurse from the rehabilitation health facility was made to confirm ventilation status on day 28 and their last day of mechanical ventilation.

**Pre-specified secondary outcomes, evaluated up to day 28, included:**

1) All-cause death;
2) Composite of ICU admission or all-cause death;
3) Composite of mechanical ventilation or all-cause death;
4) Major bleeding as defined by the ISTH Scientific and Standardization Committee (ISTH-SSC) recommendation;[1]
5) Red blood cell transfusion ($\geq$1 unit);
6) Transfusion of platelets, frozen plasma, prothrombin complex concentrate, cryoprecipitate and/or fibrinogen concentrate;
7) Renal replacement therapy defined as continuous renal replacement therapy {CRRT} or intermittent hemodialysis {IHD};
8) Hospital-free days alive;
9) ICU-free days alive;
10) Ventilator-free days alive;
11) Organ support-free days alive;
12) Venous thromboembolism (defined as symptomatic or incidental, suspected or confirmed via diagnostic imaging and/or electrocardiogram where appropriate recognizing that access to diagnostic imaging may have been limited due to the COVID-19 pandemic, however confirmatory testing was encouraged);
13) Arterial thromboembolism (defined as suspected or confirmed via diagnostic imaging and/or electrocardiogram where appropriate recognizing that access to diagnostic imaging may have been limited due to the COVID-19 pandemic, however confirmatory testing was encouraged);
14) Heparin induced thrombocytopenia;
15) D-dimer at day 2+/- 24 hours.

In addition, the following pre-specified components of the primary composite outcome were pre-specified in the statistical analysis plan, but not in the protocol:

16) Proportion of subjects with ICU admission;
17) Proportion of subjects with the composite of invasive or non-invasive mechanical ventilation;
18) Proportion of subjects with invasive mechanical ventilation.

Outcome measures were obtained from participants' hospital medical records and where applicable through a telephone follow-up. The use of bi-level positive airway pressure (BIPAP) or continuous positive airway pressure (CPAP) at night or when sleeping for sleep apnea was not considered non-invasive mechanical ventilation or organ support for the purpose of this trial.

**3.4 Pre-specified Outcome Definition for Organ-support Free Days**

Defined as the number of days that a patient was alive and free of organ support through 28 days after trial entry. Organ support was defined as receipt of non-invasive mechanical ventilation, high flow nasal cannula oxygen, invasive mechanical ventilation, or vasopressor therapy.

- Non-invasive mechanical ventilation was defined as BIPAP or CPAP when used for acute respiratory support.
- High Flow Nasal Cannula Oxygen was defined as receiving ≥ 30 l/min flow at FiO2 ≥ 40%.
- Invasive mechanical ventilation was defined as positive pressure ventilation through endotracheal tube or tracheostomy.
- Vasopressor support included the infusion of any vasoactive or inotropic medication.
- A patient must have been extubated and not receiving mechanical ventilation for at least 2 days before being considered free of mechanical ventilation. If a patient was extubated and re-intubated and placed back on mechanical ventilation within 1 or 2 days, the patient was considered to be on mechanical ventilation during those 1 or 2 days before re-intubation.
- Any patient who died during the acute hospital stay was assigned 28 Day Organ-Support Free Days of –1.
- If there was intervening time in which a patient was free of organ support, but went back on organ support, the intervening time did not count toward the organ support free days endpoint. Only time before organ support and after the last use of organ support was counted as "free days".
- If a patient was discharged alive without mechanical ventilation prior to Day 28, the patient was assumed to be free of organ support after hospital discharge for the remainder of the 28 days.
- If a patient was discharged alive on mechanical ventilation (invasive or non-invasive) prior to day 28, a call to the patient or a doctor/nurse from the rehabilitation health facility was made to confirm ventilation status on day 28 and their last day of mechanical ventilation.

**3.5 Sample Size Considerations**

462 patients (231 per group) were needed to detect a 15% risk difference, from 50% in the control group to 35% in the experimental group, with power of 90% at a two-sided alpha of 0.048.[2–4] No attrition was expected. This calculation took two interim analyses into account. There was no inflation to account for losses to follow-up because we expected these to be very infrequent, and given the nature of the trial, included patients, and outcomes, we concluded an absence of the primary outcome in patients discharged alive from hospital before 28 days with missing outcome data at day 28.

**3.6 Extended Description of Statistical Methods**

**Statistical Analysis**

Primary analyses were by the intention-to-treat population of all randomized patients in accordance with the allocated intervention. We conducted a chi-square test to derive a two-sided p-value for the main analysis of the primary outcome. We used logistic regression to derive odds ratios with 95% confidence intervals. In addition, we derived differences in proportions and 95% confidence interval from logistic regression using the observed risk of the primary outcome in the control group,[5] and from a binomial model with identity link.

We conducted subgroup analyses accompanied by tests of interaction for the following variables: age, sex, BMI, time from COVID-19 symptom onset, diabetes mellitus, coronary artery disease, hypertension and race/ethnicity. Logistic regression and linear regression were used to analyse binary and continuous secondary outcomes after adjustment for age (used for stratification of randomization).

Secondary outcomes were exploratory and were not adjusted for multiple comparisons. A per-protocol analysis of the primary outcome was restricted to the per-protocol population of participants, defined as those who received experimental or control intervention as allocated during the first 48 hours after randomization.

If an outcome was missing in more than 5% of the patients, in addition to the pre-planned strategy of assuming no outcome if patients were discharged alive from hospital before 28 days, a complete case analysis, an inverse probability weighted analysis and multiple imputation on outcome was also conducted.

The statistical analysis plan was finalized prior to study closure without prior inspection of the data. All analyses were conducted in R version 3.6.2 and/or Stata version 15.1, or higher.

**Interim Analysis**
Interim analyses were done when approximately 25% and 75% of the originally planned number of participants reached determination of the primary outcome. A group sequential design was employed that applied a one-sided boundary. The boundary was based on a Hwang-Shih-DeCani spending function for efficacy. When approximately 75% of the originally planned number of participants reached determination of the primary endpoint, we performed a conditional power analysis.

If the conditional power given the accumulated data was <30% and there was robust evidence of harm – either a relevant increase in the risk of major bleeding in the experimental group and the lower limit of the 95% confidence interval for major bleeding excluded 5% on an absolute risk difference scale; or a relevant increase in the risk of all-cause death in the experimental group and the lower limit of the 95% confidence interval for death excluded 1% on an absolute risk difference scale - the protocol called for a non-binding recommendation to stop the trial by the Data and Safety Monitoring Board (DSMB). If the conditional power was <30%, but there was no robust evidence of harm, the protocol called for completing recruitment as planned. The rationale for this approach was that prevention of death (a component of the primary outcome) overrides short-term safety. If major bleeds led to bleeding related deaths to such an extent that no mortality benefit was likely to be realized, the trial would have been stopped. If the conditional power was ≥30 and <60%, the protocol called for completing recruitment as planned. If the conditional power was ≥60% and <80%, the protocol called for a non-binding recommendation to increase the sample size to achieve 80% power, if deemed feasible from a recruitment perspective. If the conditional power was ≥80%, the protocol called for completing recruitment as planned, provided that the interim analysis against the one-sided boundary for efficacy was negative.

**3.7 Adjudication**
Outcomes were independently and blindly adjudicated by two clinical content experts for the English language source documentation, and two clinical content experts for the Portuguese source documentation from the Brazilian site.

The adjudicators reviewed de-identified and treatment allocation redacted, source documentation (e.g. clinical notes, discharge summary, diagnostic imaging, laboratory tests, autopsy reports etc.) to confirm the presence of clinical events specified in the protocol, and the date of occurrence for the following:

1) ISTH-defined major bleeding
2) Heparin induced thrombocytopenia (HIT)
3) Venous thromboembolism
4) Arterial thromboembolism
5) Mechanical ventilation, including whether invasive or non-invasive
6) Intensive care unit admission
7) Death, including cause of death

Each clinical event was reviewed in duplicate by two independent adjudicators who determined whether the event met the pre-specified criteria (per definitions in the protocol). The events were classified as a “Definite Event", "Probable Event" or “Not an Event” (see adjudication table from the adjudication manual below). The final adjudication result was based on consensus. If there was disagreement between the two adjudicators the medical monitors broke the tie. Additional source documentation could be requested.

**Adjudication Checklist:**
**<u>Adjudicator to complete:</u>**

☐ **Definite Event**

☐ **Probable Event**

☐ **Not an Event**

**If major hemorrhage, indicate which criterion of the ISTH definition was met:**

1. ☐ Fatal bleeding, and/or

2. ☐ Symptomatic bleeding in critical area or organ (such as intracranial, intraspinal, intraocular, retroperitoneal, intraarticular or pericardial, or intramuscular with compartment syndrome, and/or

3. ☐ Bleeding causing a fall of hemoglobin level of 20g/L or more, or leading to transfusion of two or more units of whole blood or red cells.

**If HIT,** indicate if laboratory test proven ☐ Yes / ☐ No and if accompanied by a thrombotic event ☐Yes / ☐ No

**If ATE,** indicate if suspected ATE ☐ <u>OR</u> diagnostically confirmed ATE ☐

**If ATE,** indicate type: ☐ ischemic stroke <u>OR</u> ☐ MI, <u>OR</u> ☐ limb ischemia <u>OR</u> ☐other _____________
_____________________________________________________________________________________

**If VTE,** indicate if suspected VTE ☐ <u>OR</u> diagnostically confirmed VTE ☐

**If VTE,** indicate if symptomatic VTE ☐ <u>OR</u> asymptomatic VTE ☐ <u>OR</u> unclear ☐

**If VTE,** indicate type: ☐ PE <u>OR</u> ☐ DVT <u>OR</u> ☐ splanchnic VT <u>OR</u> ☐ cerebral VT <u>OR</u> ☐ other ________
_____________________________________________________________________________________

If PE: indicate if ☐ segmental/beyond <u>OR</u> ☐ subsegmental
If DVT: indicate if ☐ proximal (above knee) <u>OR</u> ☐ distal (below knee)

**If ICU admission,** ☐ Yes, patient was admitted <u>OR</u> ☐ No, patient was not admitted <u>OR</u> ☐ unclear
If yes, indicate rationale for transfer to ICU __________________________________________
______________________________________________________________________________ <u>OR</u> ☐ unclear

**If mechanically ventilated,** indicate if non-invasive ☐ <u>OR</u> invasive ☐

**If patient died,** indicate cause of death: _____________________________________ <u>OR</u> ☐unclear

**Adjudicator signature + date: _____________________________**

**Adjudication Manual**

| EVENT TYPE | INFORMATION NEEDED | DOCUMENTS REQUIRED (EXAMPLES) |
|---|---|---|
| **Death** | ☐ Date of death<br>☐ Cause of death | Medical notes, death certificate, discharge summary |
| **ICU admission** | ☐ Date of transfer to ICU<br>☐ Rationale for transfer | Medical notes, medical orders |
| **Invasive Mechanical ventilation** | ☐ Stat date of invasive mechanical ventilation (i.e. endotracheal intubation with mechanical ventilation)<br>☐ Rationale for invasive mechanical ventilation (e.g. hypoxic respiratory failure, airway protection due to compromised neurologic status)<br>☐ Max FiO2 (e.g. 0.80 or 80%)<br>☐ Stop date (if applicable) | Medical notes, medical orders, respiratory therapy notes<br>Diagnostic imaging: Chest Xray indicating placement of endotracheal tube |
| **Noninvasive Mechanical Ventilation (Positive pressure ventilation)** | ☐ Start date<br>☐ BIPAP or CPAP<br>☐ Rationale for noninvasive mechanical ventilation (e.g. hypoxic respiratory failure)<br>☐ Max FiO2 (e.g. 0.80 or 80%)<br>☐ Stop date (if applicable) | Medical notes, medical orders, respiratory therapy notes, nursing notes |
| **Major bleeding*** | ☐ Date of onset<br>☐ Date of resolution (if applicable)<br>☐ Transfusion (# of red cell units and date transfused)<br>☐ Location of bleed in critical area or organ (e.g. intracranial, intraspinal, intraocular, retroperitoneal, intraarticular, pericardial, or intramuscular with compartment syndrome)<br>☐ Fatal bleeding (yes or no)<br>☐ Fall in hemoglobin by ≥ 20 g/L | Medical notes, medical orders, nursing notes (e.g. transfusion order/nursing documentation of transfusion administration),surgical note (if applicable)<br>Lab result: fall in hemoglobin by >/= 20 g/L (if applicable)<br>Diagnostic imaging reports (if available): CT, MRI, Ultrasound report |
| **Heparin-induced thrombocytopenia** | ☐ Date of onset<br>☐ Date of resolution (if applicable)<br>☐ Laboratory confirmation of heparin-induced thrombocytopenia<br>☐ Evidence of secondary thromboembolism (if applicable)<br>☐ Treatment given (medication order) | Medical notes, medical orders<br>Lab results:<br>- 5-day trend of platelet count **AND**<br>- Immunologic based assay (ELISA or LIA) assay evaluating for heparin-PF4 antibodies and/or Serotonin release assay<br>Diagnostic imaging (per venous thromboembolism and arterial thromboembolism categories below if patient experienced secondary thromboembolism) |

| | | |
|---|---|---|
| **Venous thromboembolism** | ☐ Date of onset<br>☐ Date of resolution (if applicable)<br>☐ Type (PE, DVT, splanchnic vein thrombosis, cerebral vein thrombosis, or other)<br>o If PE: segmental/beyond or subsegmental<br>o If DVT: distal (below knee) or proximal (above knee)<br>☐ Symptomatic or asymptomatic<br>☐ Suspected or confirmed<br>☐ Treatment given (medication order) | Medical notes, medical orders<br>Lab result: D-dimer (if available) from date of onset<br>Diagnostic imaging reports (if available): CT, Doppler Ultrasound, MRI, ventilation/perfusion lung scan |
| **Arterial thromboembolism** | ☐ Date of onset<br>☐ Date of resolution (if applicable)<br>☐ Type (ischaemic stroke, MI, or limb ischemia, other)<br>☐ Suspected or confirmed<br>☐ Treatment given (medication order) | Medical notes, medical orders<br>Lab results (if available) from date of onset:<br>- Troponin<br>- Lactate (venous or arterial)<br><br>Diagnostic imaging reports (if available): Doppler ultrasound, ultrasound, angiogram, ECG, CT, MRI, echocardiogram |
| **Organ Support via High-Flow Nasal Cannula** | ☐ Start date<br>☐ Stop date (if applicable)<br>☐ Max O2 flow rate (should be >30 L/min to qualify, per our trial definition, as HFNC)<br>☐ Max FiO2 value (e.g. 0.80 or 80%) | Medical notes, respiratory therapy notes, nursing notes |
| **Organ Support via Vasopressor/Inotropic therapy** | ☐ Start date<br>☐ Stop date (if applicable)<br>☐ Vasopressor/inotrope examples:<br>▪ Norepinephrine (Levophed, Levo)<br>▪ Epinephrine (Epi)<br>▪ Vasopressin (Vaso)<br>▪ Dopamine (Dop)<br>▪ Dobutamine (Dobu) | Medical notes, nursing notes, medical orders |

*Major bleeding defined by ISTH Scientific and Standardization Committee (ISTH-SSC) recommendation:

1) Fatal bleeding, and/or
2) symptomatic bleeding in critical area or organ (such as intracranial, intraspinal, intraocular, retroperitoneal, intraarticular or pericardial, or intramuscular with compartment syndrome, and/or
3) bleeding causing a fall of hemoglobin level of 20g/L or more, or leading to transfusion of two or more units of whole blood or red cells.

#### 3.8 Data and Safety Monitoring Board

The DSMB acted in an advisory capacity to the principal investigators to monitor participant safety, data quality and evaluate the progress of the trial. The DSMB was composed of a biostatistician, a hematologist, a general internist and an intensive care specialist. The four members were not study investigators. The DSMB convened meetings at the formal interim analyses mentioned above (section 3.6), and also when approximately 10% and 50% of the originally planned number of participants reached determination of the primary outcome. When 10%, 25% and 50% of the originally planned number of participants reached determination of the primary outcome, this recommendation was at the discretion of the DSMB. When approximately 75% of the originally planned number of participants reached determination of the primary outcome, the DSMB was required to also take into account the conditional power when doing the safety review. The recommendations were made by a formal majority vote based on safety concerns as evidenced by statistical and clinical judgment, progress of the trial including data quality and accrual/retention, and new scientific or therapeutic developments that may have an impact on the safety. The DSMB was immediately informed of any serious adverse events (SAEs) which were potentially study drug related. Moreover, the DSMB chair was notified within 24 hours of any major bleed or occurrence of heparin induced thrombocytopenia. The Data Management and Coordination Center (DMCC) was responsible for the data analysis and the DMCC statistician provided the interface with DSMB members. The DSMB received reports of enrollment and events, including events as reported by the site and had full access to the data.

**3.9 Trial Administration**

Protocols in English (NCT04362085) and Portuguese (NCT04444700) were harmonized in all aspects, except for an initial material difference in eligibility criteria: between July 3, 2020, and October 22, 2020, patients in the single Brazilian site of the trial were eligible if they had normal D-Dimer levels, but an oxygen saturation of ≤93% on room air. This led to the initial inclusion of 11 patients in Brazil with normal D-Dimer levels at baseline. These patients were included in the intention-to-treat analysis. From October 23, 2020 onwards, protocols were fully harmonized, requiring elevated D-Dimer levels at baseline in all patients. Data from all sites, including the site in Brazil, were entered in a common database and managed centrally by the trial's data coordination centre (Applied Health Research Centre, St. Michael's Hospital, University of Toronto) according to a single set of standard operating procedures.

The original protocol of RAPID BRAZIL initially approved on June 17, 2020 by the Institutional Research Ethics Board and Brazilian National Research Ethics Commission is version 3.0. All subsequent versions were anchored from version 3.0. The latest version of the protocol is version 5.2 approved on April 13, 2021.

### 4. Supplementary Figures and Tables

#### Figure S1. Trial Schematic

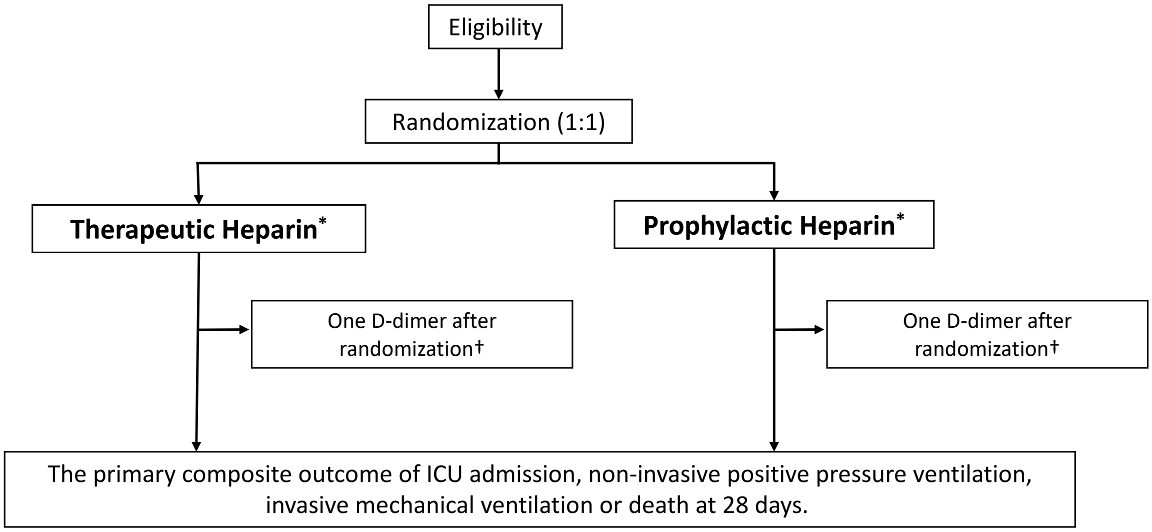

*Administered until hospital discharge, death or day 28, if the patient is admitted to the ICU or required ventilatory support, we recommended continuation of the allocated treatment as long as the treating physician was in agreement.
†A single D-dimer test (if not collected through standard of care) on day 2 after randomization (±24 hours) was collected for participants in both study arms (considering the day of randomization as day 1).

**Table S1. Duration and Type of Heparin Used**

| Heparin | Therapeutic Heparin (N=228) | Prophylactic Heparin (N=237) |
|---|---|---|
| | *no. of patients (%)* | |
| Duration of anticoagulation (days)* | | |
| Mean (SD) | 6.5 (5.4) | 6.3 (5.4) |
| Median (IQR) | 6.0 [3.0, 8.0] | 5.0 [3.0, 8.0] |
| Dalteparin | 25 (11.0) | 25 (10.5) |
| Enoxaparin | 188 (82.5) | 183 (77.2) |
| Fondaparinux | 0 (0.0) | 1 (0.4) |
| Tinzaparin | 11 (4.8) | 13 (5.5) |
| Unfractionated heparin | 3 (1.3) | 14 (5.9) |

SD: standard deviation; IQR: interquartile range.

*Data on the duration of anticoagulation were missing for 4 patients in the therapeutic heparin group and 5 patients in the prophylactic heparin group .

**Table S2. Concomitant Treatments Received**

| | Therapeutic Heparin | | Prophylactic Heparin | |
|---|---|---|---|---|
| | No Death (N=224) | Death (N=4) | No Death (N=219) | Death (N=18) |
| | *no. of patients (%)* | | | |
| Systemic Corticosteroid | 172 (76.8) | 3 (75.0) | 167 (76.3) | 14 (77.8) |
| Remdesivir | 30 (13.4) | 1 (25.0) | 27 (12.3) | 3 (16.7) |
| Tocilizumab | 10 (4.5) | 0 (0.0) | 11 (5.0) | 2 (11.1) |

Treatments received over course of study duration, pre- and post-randomization combined.

**Table S3. Primary Cause of Death**

| Cause | Therapeutic Heparin (N=228) | Prophylactic Heparin (N=237) |
|---|---|---|
| | *no. of patients (%)* | |
| Hypoxic respiratory failure | 4 (100.0) | 13 (72.2) |
| Multi-system organ failure | 0 (0.0) | 5 (27.8) |

There were no cases of sudden, unexplained death.

**Table S4. Thromboembolism**

| Event | Therapeutic Heparin§ (N=228) | Prophylactic Heparin (N=237) |
|---|---|---|
| | *no. of patients (%)* | |
| Venous | | |
| Deep vein thrombosis† | 1 (0.4) | 2 (0.8) |
| Pulmonary embolism* | 1 (0.4) | 5 (2.1) |
| Arterial | | |
| Myocardial infarction | 0 (0.0) | 1 (0.4) |

†1 patient in the therapeutic heparin group (symptomatic, diagnostically confirmed, proximal deep vein), 2 patients in the prophylactic heparin group (1 symptomatic, could not be definitively confirmed as diagnostic imaging not done during acute symptomatic period; 1 incidental, diagnostically confirmed, proximal deep vein).
*1 patient in the therapeutic heparin group (symptomatic, diagnostically confirmed, segmental pulmonary artery or beyond), 5 patients in the prophylactic heparin group (all symptomatic, all diagnostically confirmed, 4 segmental pulmonary artery or beyond, 1 sub-segmental pulmonary artery).

**Figure S2. Subgroup Analysis of the Primary Outcome**

| Subgroup | Therapeutic Heparin | Prophylactic Heparin | Odds Ratio (95% CI) | P for Interaction |
|---|---|---|---|---|
| All Patients | 37 (16.2) | 52 (21.9) | 0.69 (0.43 to 1.10) | |
| Age | | | | 0.59 |
| <=65 | 20 (8.8) | 25 (10.5) | 0.78 (0.41 to 1.48) | |
| >65 | 17 (7.5) | 27 (11.4) | 0.60 (0.30 to 1.21) | |
| Sex | | | | 0.73 |
| Male | 26 (11.4) | 37 (15.6) | 0.75 (0.42 to 1.34) | |
| Female | 11 (4.8) | 15 (6.3) | 0.63 (0.27 to 1.46) | |
| Race/ethnicity | | | | 0.47 |
| White | 26 (11.8) | 41 (17.5) | 0.57 (0.33 to 0.99) | |
| Asian | 4 (1.8) | 5 (2.1) | 1.15 (0.28 to 4.76) | |
| Black or African American | 3 (1.4) | 2 (0.9) | 2.10 (0.31 to 14.22) | |
| Hispanic or Latino | 2 (0.9) | 3 (1.3) | 0.39 (0.05 to 2.94) | |
| BMI | | | | 0.84 |
| <=30 | 22 (9.9) | 30 (12.9) | 0.69 (0.38 to 1.28) | |
| >30 | 13 (5.9) | 21 (9) | 0.63 (0.29 to 1.34) | |
| Hypertension | | | | 0.30 |
| Yes | 18 (7.9) | 16 (6.8) | 0.57 (0.30 to 1.05) | |
| No | 19 (8.3) | 35 (14.8) | 1.00 (0.47 to 2.11) | |
| Diabetes mellitus | | | | 0.17 |
| Yes | 3 (1.3) | 7 (3) | 1.06 (0.49 to 2.26) | |
| No | 32 (14) | 44 (18.6) | 0.53 (0.29 to 0.98) | |
| Coronary artery disease | | | | 0.42 |
| Yes | 21 (9.2) | 35 (14.8) | 0.36 (0.07 to 1.76) | |
| No | 16 (7) | 16 (6.8) | 0.72 (0.43 to 1.19) | |
| Duration of symptoms prior to hospitalization | | | | 0.091 |
| <=7 days | 29 (13.4) | 31 (13.4) | 0.97 (0.55 to 1.74) | |
| >7 days | 8 (3.7) | 18 (7.8) | 0.39 (0.16 to 0.95) | |
| Region | | | | 0.38 |
| North America | 13 (5.7) | 23 (9.7) | 0.54 (0.25 to 1.16) | |
| Brazil | 9 (3.9) | 15 (6.3) | 0.48 (0.19 to 1.23) | |
| Middle East | 14 (6.1) | 12 (5.1) | 1.28 (0.55 to 2.97) | |
| Ireland | 1 (0.4) | 2 (0.8) | 0.50 (0.04 to 6.48) | |
| D−dimer levels | | | | 0.38 |
| <2x upper limit of normal | 14 (6.1) | 26 (11) | 0.54 (0.26 to 1.09) | |
| >=2x upper limit of normal | 23 (10.1) | 26 (11) | 0.83 (0.44 to 1.56) | |
| Use of systemic corticosteroids | | | | 0.32 |
| Yes | 29 (12.7) | 35 (14.8) | 0.80 (0.46 to 1.38) | |
| No | 8 (3.5) | 17 (7.2) | 0.46 (0.18 to 1.16) | |

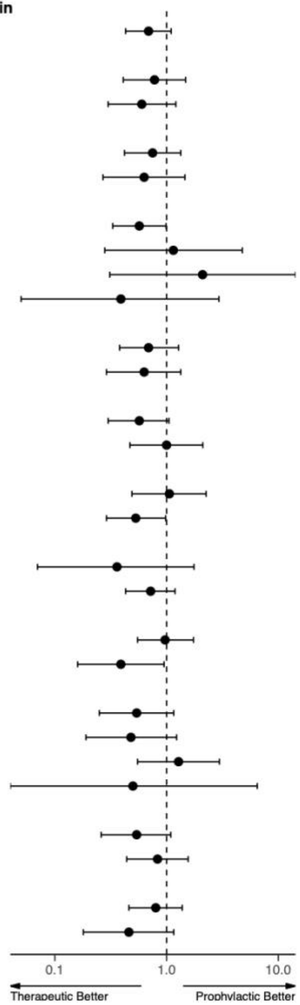

Subgroup-specific odds ratios derived from logistic regression. Point estimates are plotted as dark circles; the horizontal lines represent the 95% confidence intervals. Odds ratio less than 1.0 favours therapeutic heparin. BMI, body mass index in kg/m$^2$).

**Table S5. Per Protocol Analysis of the Primary Outcome and Its Components**

| Outcome | Therapeutic Heparin (N=216) | Prophylactic Heparin (N=227) | Odds Ratio (95% CI) |
|---|---|---|---|
| | *no. of patients (%)* | | |
| Primary Composite Outcome† | 34 (15.7) | 47 (20.7) | 0.72 (0.44, 1.17) |
| Components of the primary composite outcome | | | |
| Death from any cause | 4 (1.9) | 17 (7.5) | 0.23 (0.08, 0.71) |
| Invasive mechanical ventilation | 9 (4.2) | 13 (5.7) | 0.72 (0.30, 1.71) |
| Any mechanical ventilation° | 18 (8.3) | 22 (9.7) | 0.85 (0.44, 1.63) |
| Intensive care unit admission | 30 (13.9) | 37 (16.3) | 0.83 (0.49, 1.40) |

Per protocol analysis excluded patients who did not receive their allocated treatment during the first 48 hours after randomization.

†Defined as death, invasive mechanical ventilation, non-invasive mechanical ventilation or ICU admission.

°Invasive or non-invasive (bilevel or continuous positive airway pressure) mechanical ventilation.

**Table S6. Sensitivity Analysis 1 of the Primary Outcome and Its Components**

| Outcome | Therapeutic Heparin (N=217) | Prophylactic Heparin (N=225) | Odds Ratio (95% CI) |
|---|---|---|---|
| | *no. of patients (%)* | | |
| Primary Composite Outcome† | 37 (17.1) | 52 (23.1) | 0.68 (0.43, 1.10) |
| Components of the primary composite outcome | | | |
| Death from any cause | 4 (1.8) | 18 (8.0) | 0.22 (0.07, 0.65) |
| Invasive mechanical ventilation | 11 (5.1) | 16 (7.1) | 0.70 (0.32, 1.54) |
| Any mechanical ventilation° | 21 (9.7) | 26 (11.6) | 0.82 (0.45, 1.51) |
| Intensive care unit admission | 33 (15.2) | 42 (18.7) | 0.78 (0.47, 1.29) |

Sensitivity analysis 1 excluded patients who did not meet a component of the primary composite outcome and did not have a follow-up up to day 28; 11 patients in therapeutic heparin group and 12 patients in the prophylactic heparin group.

†Defined as death, invasive mechanical ventilation, non-invasive mechanical ventilation or ICU admission.

°Invasive or non-invasive (bilevel or continuous positive airway pressure) mechanical ventilation.

**Table S7. Sensitivity Analysis 2 of the Primary Outcome and Its Components**

| Outcome | Therapeutic Heparin (N=222) | Prophylactic Heparin (N=231) | Odds Ratio (95% CI) |
|---|---|---|---|
| | *no. of patients (%)* | | |
| Primary Composite Outcome† | 36 (16.2) | 48 (20.8) | 0.74 (0.46, 1.19) |
| Components of the primary composite outcome | | | |
| Death from any cause | 4 (1.8) | 17 (7.4) | 0.23 (0.08, 0.70) |
| Invasive mechanical ventilation | 10 (4.5) | 14 (6.1) | 0.73 (0.32, 1.69) |
| Any mechanical ventilation° | 20 (9.0) | 23 (10.0) | 0.90 (0.48, 1.68) |
| Intensive care unit admission | 32 (14.4) | 39 (16.9) | 0.83 (0.50, 1.38) |

Sensitivity analysis 2 excluded those who did not satisfy all eligibility criteria (i.e. those with a negative d-dimer; 6 patients in the therapeutic heparin group and 5 in the prophylactic heparin group).

†Defined as death, invasive mechanical ventilation, non-invasive mechanical ventilation or ICU admission.

°Invasive or non-invasive (bilevel or continuous positive airway pressure) mechanical ventilation.

**Table S8. Sensitivity Analysis 3 of the Primary Outcome and Its Components**

| Outcome | Therapeutic Heparin (N=211) | Prophylactic Heparin (N=219) | Odds Ratio (95% CI) |
|---|---|---|---|
| | *no. of patients (%)* | | |
| Primary Composite Outcome† | 36 (17.1) | 48 (21.9) | 0.73 (0.45, 1.19) |
| Components of the primary composite outcome | | | |
| Death from any cause | 4 (1.9) | 17 (7.8) | 0.23 (0.08, 0.70) |
| Invasive mechanical ventilation | 10 (4.7) | 14 (6.4) | 0.73 (0.32, 1.68) |
| Any mechanical ventilation° | 20 (9.5) | 23 (10.5) | 0.89 (0.47, 1.68) |
| Intensive care unit admission | 32 (15.2) | 39 (17.8) | 0.83 (0.49, 1.38) |

Sensitivity analysis 3 excluded patients who did not meet a component of the primary composite outcome, did not have a follow-up up to day 28 and those who did not satisfy all eligibility criteria; 17 patients in the therapeutic heparin group and 18 patients in the prophylactic heparin group.

†Defined as death, invasive mechanical ventilation, non-invasive mechanical ventilation or ICU admission.

°Invasive or non-invasive (bilevel or continuous positive airway pressure) mechanical ventilation.

**Table S9. Intention-to-Treat Analysis of the Primary Outcome and Its Components Adjusted for Age**

| Outcome | Therapeutic Heparin (N=228) | Prophylactic Heparin (N=237) | Odds Ratio (95% CI) |
|---|---|---|---|
| | *no. of patients (%)* | | |
| Primary Composite Outcome† | 37 (16.2) | 52 (21.9) | 0.68 (0.42, 1.08) |
| Components of the primary composite outcome | | | |
| Death from any cause | 4 (1.8) | 18 (7.6) | 0.19 (0.06, 0.61) |
| Invasive mechanical ventilation | 11 (4.8) | 16 (6.8) | 0.69 (0.31, 1.53) |
| Any mechanical ventilation° | 21 (9.2) | 26 (11.0) | 0.82 (0.45, 1.50) |
| Intensive care unit admission | 33 (14.5) | 42 (17.7) | 0.78 (0.47, 1.29) |

Intention-to-treat analysis of the primary outcome and its components adjusted for age taking into account that randomization was stratified by age.

†Defined as death, invasive mechanical ventilation, non-invasive mechanical ventilation or ICU admission.

°Invasive or non-invasive (bilevel or continuous positive airway pressure) mechanical ventilation.

**Table S10. Intention-to-Treat Analysis of the Primary Outcome with Time-by-Treatment Interaction**

| Analysis | Therapeutic Heparin (N=228) | Prophylactic Heparin (N=237) | Odds Ratio (95% CI) |
|---|---|---|---|
| | *no. of patients (%)* | | |
| Primary Analysis | 37 (16.2) | 52 (21.9) | 0.69 (0.43, 1.10) |
| Analysis adjusted for time | 37 (16.2) | 52 (21.9) | 0.69 (0.43, 1.10) |

Intention-to-treat analysis of the primary outcome according to primary analysis and adjusted for time, including a time-by-treatment interaction.

Primary outcome defined as death, invasive mechanical ventilation, non-invasive mechanical ventilation or ICU admission.

To address changes in co-interventions over time due to emerging evidence from Covid-19 clinical trials, a logistic regression model was used to fit a time by treatment interaction where time was days since first randomized subject. Time was modelled with a restricted cubic spline having 3 knots. Three knots were chosen because of the modest number of events.

The model with splines and interactions revealed little evidence for an interaction ($p = 0.85$) or non-linearity ($p = 0.95$). Given these results a linear additive model was fit to estimate the time adjusted treatment effect. In this model there was strong evidence of a time effect ($p=0.0086$) while the evidence for a treatment effect was identical to the unadjusted analysis (p for treatment effect=0.12).

**Table S11. Intention-to-Treat Analysis of the Primary Outcome Estimating Risk Differences**

| **Analysis** | **Therapeutic Heparin (N=228)** | **Prophylactic Heparin (N=237)** | **Risk Difference (95% CI)** |
|---|---|---|---|
| | *no. of patients (%)* | | |
| Estimated from logistic regression | 37 (16.2) | 52 (21.9) | -5.7% (-11.2%, 1.7%) |
| Estimated from binomial model | 37 (16.2) | 52 (21.9) | -5.7% (-12.9%, 1.4%) |

Intention-to-treat analysis of the primary outcome estimating risk differences from logistic regression and a binomial model with identity link.
Primary outcome defined as death, invasive mechanical ventilation, non-invasive mechanical ventilation or ICU admission.

The primary outcome was reanalyzed with a binary model and identity link to estimate the absolute risk difference. This analysis yielded nearly identical results to the logistic regression and risk difference estimated from that model. The evidence for a treatment effect was similar to the evidence based on logistic regression (p for treatment effect=0.12).

**Table S12. Sensitivity Analyses of D-Dimer Levels at Day 2**

| **Analysis** | **Ratio of Geometric Means (95% CI)** |
|---|---|
| Primary analysis (complete case) | 0.88 (0.78, 0.99) |
| Inverse probability weighted analysis | 0.87 (0.78, 0.98) |
| Multiple Imputation | 0.91 (0.79, 1.04) |

D-dimer levels at day 2±24 hours post-randomization were missing for 66 (29.0%) in the therapeutic heparin group and 64 (27.0%) in the prophylactic heparin groups. As pre-specified, we therefore used an inverse probability weighted analysis and multiple imputation to derive ratios of geometric means.
Ratio of geometric means of D-dimer level x ULN of day 2±24h post-randomization, adjusted for baseline geometric means of D-dimer levels x ULN using analysis of covariance. SD for the natural logarithm of D-dimer levels x ULN.

**Table S13. ISTH Major Bleeding Events**

| | Randomized treatment allocation | Fatal bleeding | Symptomatic bleeding in critical area or organ* | Bleeding causing a fall of hemoglobin of 20g/L or more | Bleeding leading to transfusion of two or more units of whole blood or red cells | Relatedness† | Concomitant medications |
|---|---|---|---|---|---|---|---|
| Patient 1 | Therapeutic Heparin | No | Intramuscular | Yes | Yes | Not related | Systemic corticosteroid |
| Patient 2 | Therapeutic Heparin | No | No | Yes | Yes | Unlikely | Systemic corticosteroid, Antiplatelet agent |
| Patient 3 | Prophylactic Heparin | No | No | Yes | Yes | Unlikely | Systemic corticosteroid |
| Patient 4 | Prophylactic Heparin | No | No | Yes | Yes | Unlikely | Systemic corticosteroid, Antiplatelet agent |
| Patient 5 | Prophylactic Heparin | No | Retroperitoneal | Yes | Yes | Probable | Antiplatelet agent |
| Patient 6 | Prophylactic Heparin | No | No | Yes | Yes | Unlikely | Systemic corticosteroid |

Major bleeding defined by the International Society on Thrombosis and Haemostasis (ISTH) Scientific and Standardization Committee.[1]
*All non-critical area/organ bleeding events were gastrointestinal in origin.
†Relatedness was independently and blindly adjudicated.

**Table S14. Bleeding Events by Concomitant Treatments Received**

| | Therapeutic Heparin | Prophylactic Heparin |
|---|---|---|
| | *no. of patients (%)* | |
| Major Bleeding | 2 | 4 |
| Systemic corticosteroid only | 2 (100.0) | 2 (50.0) |
| Antiplatelet agent only | 0 (0.0) | 1 (25.0) |
| Antiplatelet and systemic corticosteroid | 0 (0.0) | 1 (25.0) |
| No antiplatelet and systemic corticosteroid | 0 (0.0) | 0 (0.0) |
| No major bleeding | 226 | 233 |
| Systemic corticosteroid only | 125 (55.3) | 129 (55.4) |
| Antiplatelet agent only | 8 (3.5) | 13 (5.6) |
| Antiplatelet and systemic corticosteroid | 25 (11.1) | 24 (10.3) |
| No antiplatelet and systemic corticosteroid | 68 (30.1) | 67 (28.8) |

Major bleeding defined by the International Society on Thrombosis and Haemostasis Scientific and Standardization Committee.[1]

**Figure S3. Analyses of the Interaction between Treatment Effect and Severity of Illness of Therapeutic Heparin versus Usual Care in Patients with Covid-19**

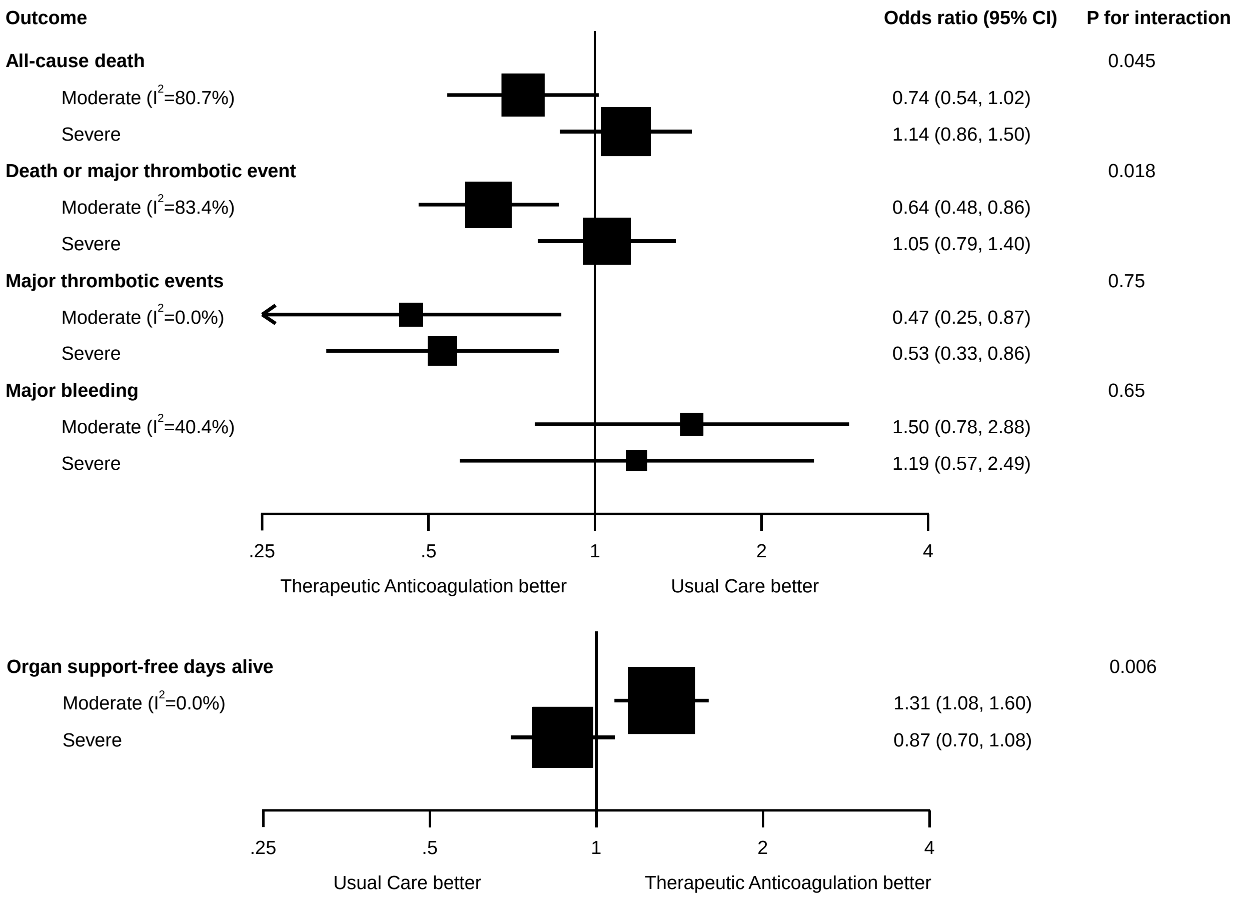

The analysis is based on Mantel-Haenszel fixed-effect meta-analyses of the RAPID trial and the multiplatform trial in moderately ill ward patients,[6] and results of the multiplatform trial in severely ill ICU patients.[7] Squares and horizontal lines show treatment effects and their 95% confidence intervals in each subgroup. The area of each square is proportional to the inverse of the variance in the subgroup. Odds ratios for organ support-free days alive are from ordinal logistic regression in all trials; death up to 28 days was assigned the worst outcome (a value of -1) in all trials. The p-values for interaction are for the comparison of treatment effects between moderately and severely ill patients and were derived from a Chi-squared test. Major thrombotic events were defined as the composite of myocardial infarction, pulmonary embolism, ischemic stroke or systemic arterial embolism; Major bleeding defined by the ISTH Scientific and Standardization Committee.[1] In accordance with the primary outcome definition of the multiplatform trials,[6,7] organ support-free days alive were calculated for an observation time of 21 days; remaining outcomes were based on an observation time of 28 days.

**Table S15. Study Drug Not Received as Allocated within the First 48 hours**

| | Therapeutic Heparin (N=228) | Prophylactic Heparin (N=237) |
|---|---|---|
| | *no. of patients (%)* | |
| Any change | 6 (2.6) | 5 (2.1) |
| Discontinued | 2 (0.9) | 1 (0.4) |
| Prophylactic | 2 (0.9) | - |
| Intermediate | 2 (0.9) | 1 (0.4) |
| Therapeutic | - | 3 (1.3) |

Study drug not received as allocated defined as not received as allocated within the first 48 hours post randomization or changed without clear clinical indication.

### 5. References

1. Schulman S, Kearon C. Definition of major bleeding in clinical investigations of antihemostatic medicinal products in non-surgical patients. Journal of Thrombosis and Haemostasis. 2005;3(4):692–4.

2. Tang N, Bai H, Chen X, Gong J, Li D, Sun Z. Anticoagulant treatment is associated with decreased mortality in severe coronavirus disease 2019 patients with coagulopathy. J Thromb Haemost. 18(5):1094–9.

3. Zhou F, Yu T, Du R, Fan G, Liu Y, Liu Z, et al. Clinical course and risk factors for mortality of adult inpatients with COVID-19 in Wuhan, China: a retrospective cohort study. The Lancet. 2020 Mar;395(10229):1054–62.

4. Sholzberg M, Tang GH, Negri E, Rahhal H, Kreuziger LB, Pompilio CE, et al. Coagulopathy of hospitalised COVID-19: A Pragmatic Randomised Controlled Trial of Therapeutic Anticoagulation versus Standard Care as a Rapid Response to the COVID-19 Pandemic (RAPID COVID COAG – RAPID Trial): A structured summary of a study protocol for a randomised controlled trial. Trials. 2021 Mar 10;22(1):202.

5. Doi SA, Furuya-Kanamori L, Xu C, Lin L, Chivese T, Thalib L. Questionable utility of the relative risk in clinical research: a call for change to practice. J Clin Epidemiol. 2020 Nov 7;

6. Lawler PR, Goligher EC, Berger JS, Neal MD, McVerry BJ, Nicolau JC, et al. Therapeutic Anticoagulation in Non-Critically Ill Patients with Covid-19. medRxiv. 2021 Jan 1;2021.05.13.21256846.

7. Goligher E, Bradbury C, McVerry BJ, Lawler PR, Berger J, Gong MN, et al. Therapeutic Anticoagulation in Critically Ill Patients with Covid-19 – Preliminary Report. medRxiv. 2021 Jan 1;2021.03.10.21252749.
